# Methods for Mediation Analysis with High-Dimensional DNA Methylation Data: Possible Choices and Comparison

**DOI:** 10.1101/2023.02.10.23285764

**Authors:** Dylan Clark-Boucher, Xiang Zhou, Jiacong Du, Yongmei Liu, Belinda L Needham, Jennifer A Smith, Bhramar Mukherjee

## Abstract

Epigenetic researchers often evaluate DNA methylation as a mediator between social/environmental exposures and disease, but modern statistical methods for jointly evaluating many mediators have not been widely adopted. We compare seven methods for high-dimensional mediation analysis with continuous outcomes through both diverse simulations and analysis of DNAm data from a large national cohort in the United States, while providing an R package for their implementation. Among the considered choices, the best-performing methods for detecting active mediators in simulations are the Bayesian sparse linear mixed model by Song et al. (2020) and high-dimensional mediation analysis by Gao et al. (2019); while the superior methods for estimating the global mediation effect are high-dimensional linear mediation analysis by Zhou et al. (2021) and principal component mediation analysis by Huang and Pan (2016). We provide guidelines for epigenetic researchers on choosing the best method in practice and offer suggestions for future methodological development.

## Introduction

In this study, we review and evaluate the available methods for performing mediation analysis when the mediators are high-dimensional DNA methylation (DNAm) measurements. DNAm is an epigenomic mechanism describing when a methyl group binds to the DNA, which occurs predominantly at cytosine-guanine dinucleotides, called “CpG sites.” DNAm has an important role in regulating gene expression across the entire genome, and is particularly impactful at CpG sites in the promoter regions of genes, where it can inhibit the binding of enzymes needed for transcription^1^.

Recent advancements in technology have made it possible to collect DNAm data on a massive scale^2^. Indeed, microarray technologies have enabled the measurement of over 850,000 CpG sites simultaneously^2^, encouraging broad research on DNAm in the etiology of disease; and studies taking advantage of these tools have identified DNAm as a risk factor in obesity^3,4^, type II diabetes^5^ and cardiovascular conditions^6,7^. At the same time, however, DNAm has also been linked to exposures such as diet^8^, smoking^9^, alcohol^10^, air pollution^11^, and socioeconomic status (SES)^12,13^, which has prompted research on whether the effects of these exposures on health outcomes could be transferred by changes in DNAm. Effect transmission of this nature is called *mediation*, and it has become popular in epigenomic research to treat DNAm as a high-dimensional mediator between environmental exposures and human disease^14^.

As an example of such an analysis, our previous work^15,16^ examined the association between low SES and glycated hemoglobin (HbA1c) in the Multi-Ethnic Study of Atherosclerosis (MESA), a United States population-based longitudinal study^17^. Indicators of SES, such as education level, are strong predictors of type II diabetes^18^, while HbA1c is an important risk factor of cardiovascular disease and a critical biomarker in type II diabetes diagnosis^19–21^. Since education level is also associated with DNAm^12,13,22^, and DNAm itself with HbA1c level^23^, we hypothesized that if low education results in greater HbA1c, part of that effect could be mediated by DNAm (Fig. 1). In the current study, we revisit this hypothesis for the purpose of illustration. Our sample from MESA has 963 individuals and includes DNAm measurements at 402,339 CpG sites, none of which we know for certain are related to education or HbA1c in advance.

**Fig. 1.**
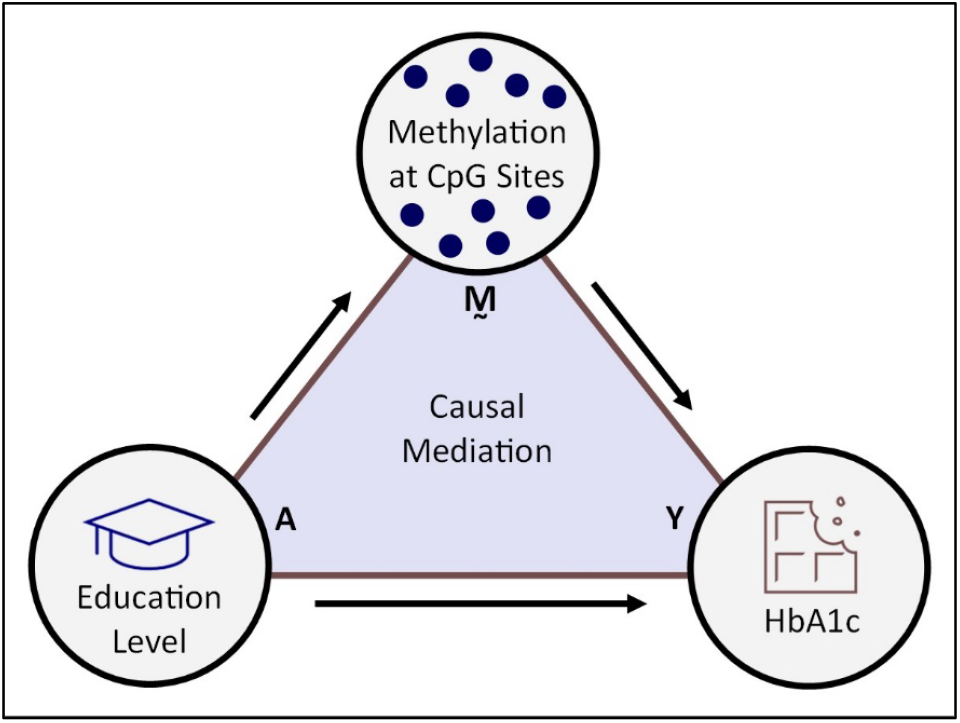
Proposed causal mechanism in which the effect of low education on HbA1c is mediated by DNAm.

The standard statistical tool for addressing such a hypothesis is mediation analysis. Formally, mediation is when an exposure, say *A*, affects an outcome, *Y*, in part through its effect on a single mediating variable *M*. When *M* is a mediator of the *A* to *Y* association, the total effect of *A* on *Y* has two components: an *indirect effect*, from *A* affecting *M* and *M* affecting *Y*, and a *direct effect*, from *A* affecting *Y* independently of *M*. In the “traditional mediation analysis” approach proposed by Baron and Kenny (1986), the associations from this mechanism could be measured by fitting a few regression models: one for the effect of *A* on *M* (the mediator model), one for the effects of *A* and *M* on *Y* (the outcome model), and sometimes a third model for the total effect of *A* on *Y, M* ignored^24–26^. The more recently developed “causal mediation analysis,” based on the counterfactual approach^27,28^, has established conditions under which the parameters of these models can be interpreted as causal effects^29^. The causal approach is more flexible when *Y* or *M* are binary and when there is *A*-*M* interaction in the outcome model^30^.

While standard examples of mediation consider only one exposure, one mediator, and one outcome^31,32^, there has been growing interest in methods for mediation that can handle many potential mediators at once. Epigenetic studies have felt this need especially, as DNAm is usually measured at several hundred thousand CpG sites with little prior knowledge of their importance. In settings such as this, a naïve strategy would be to evaluate the potential mediators one at a time, each with their own pair of models; but if the mediators are correlated this approach is inefficient, and the resulting estimates are potentially biased due to confounding from the excluded co-mediators^31^. Instead, so that we leverage these correlations rather than ignore them, the preferred approach is to assess the mediators jointly, in a single multivariable model. Although several methods for fitting such a model have been presented in the literature, none of them are widely used in analyzing DNAm data, a sign that epigenetic research is still catching up to recent developments in mediation analysis with high-dimensional mediators.

Our study aims to bridge this gap and guide researchers in epigenetics to use state of the art methods for mediation analysis with high-dimensional mediators. Despite the recent methodological developments, there are no clear-cut standards for which methods should be applied in which circumstances, making it difficult to select the best-suited method for an analysis in advance. While our prior research examined methods for large scale single-mediator hypotheses^31^, there is no such work for methods that can incorporate many potential mediators at once. Our study addresses this question first with an extensive simulation study, directly comparing the performance of sevesn different methods for mediation with high-dimensional mediators across a spectrum of settings. Along with metrics related to identification of key mediators and estimation of mediation effect, we include a computation time comparison to evaluate the scalability of the methods to large datasets. Next, to assess the utility of these methods on real data, we apply the same seven methods—plus two additional methods adapted from them—on the data from MESA to evaluate the mediating role of DNAm in the association between low education level and HbA1c. Our study is the first to address this critical gap in the epigenetic mediation literature, both by providing clarity on the methods available and by assessing their strengths and weaknesses under different settings. Moreover, although the analysis is centered around DNAm, the methods we deploy are not specific to epigenetics, and our results and guidelines should be similarly useful for researchers studying high-dimensional mediation problems in other fields. We include, supplementary to our study, an R package for implementing the methods, called “hdmed,” so that researchers have access to a centralized resource they can draw from in their own high-dimensional mediation analyses.

## Notations and General Framework

Before proceeding, it will be useful to provide an overview of the relevant mediation model and to summarize the types of methods which have become available. To begin, suppose we have a dataset of *n* individuals: an exposure *A*_*i*_, a continuous outcome *Y*_*i*_, and continuous mediators ***M***_*i*_ measured for the *i*^th^ person, *i* varying from 1 to *n*. We write ***M***_*i*_ in bold to indicate its status as a vector—in this case, a set of *p* mediators ***M***_*i*_^(*j*)^, *j* varying from 1 to *p*. Let ***C***_*i*_ be a vector of *q* covariates. When *p* is greater than 1, we can use the regression models

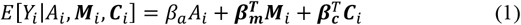

and

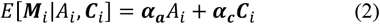

to estimate the mediating role of ***M***_*i*_ in the causal pathway between the exposure and outcome^33^. Model (1) is the outcome model and model (2) is the mediator model. In model (1), ***β***_***m***_ is a *p*-vector in which the *j*^th^ component, **(*β***_***m***_)_*j*_, is the linear association of *j*^th^ mediator with *Y*_*i*_ adjusting for the other variables; while *β*_*a*_ is the association between *A*_*i*_ and *Y*_*i*_ adjusting for mediators and covariates. In model (2), ***α***_***a***_ is a *p*-vector of the associations between the exposure and each mediator, **(*α***_***a***_)_*j*_; and ***α***_***c***_ is a matrix with the mediator-covariate associations. Also note that in model (1), we have assumed there is no interaction between *A*_*i*_ and ***M***_*i*_, which is beyond the scope of our present study.

The parameters of these models underly the causal effects of interest. Under certain assumptions^27,33^, the direct effect of *A*_*i*_ on *Y*_*i*_ is *β*_*a*_, the global indirect effect (or global mediation effect) of *A*_*i*_ on *Y*_*i*_ through ***M***_*i*_ is ***α***_***a***_ ^*T*^***β***_***m***_, and the total effect of *A*_*i*_ on *Y*_*i*_ is *β*_*a*_ *+* ***α***_***a***_^*T*^***β***_***m***_. Another quantity of interest is the proportion mediated, defined as the ratio of the global indirect effect to the total effect, which measures the degree to which the *A*_*i*_ to *Y*_*i*_ pathway is mediated by ***M***_i_. We may also seek to measure the product terms **(*α***_***a***_)_*j*_**(*β***_***m***_)_*j*_, which measure the contribution of the *j*^th^ mediator to the global indirect effect, since summing these for *j* from 1 to *p* yields ***α***_***a***_^*T*^***β***_***m***_. However, we emphasize that **(*α***_***a***_)_*j*_**(*β***_***m***_)_*j*_ cannot be interpreted as a causal effect through the *j*^th^ mediator on its own, since we have made no assumptions about the causal ordering of the mediators and can only formally treat them as a joint system. Instead, we call **(*α***_***a***_)_*j*_**(*β***_***m***_)_*j*_ the *mediation contribution*, and describe the *j*^th^ mediator as *active* if its contribution is not zero.

If the potential mediators are uncorrelated, conditional on the exposure and covariates, or if *p* is reasonably small relative to *n*, then it is trivial to fit the above models using linear regression. However, if the mediators are correlated and *p* is large, the estimates from model (1) may have extremely high variance; and if *p* is so large as to exceed *n*, the linear regression model cannot even be fitted. These concerns are relevant to us because DNAm measurements tend to be correlated, while the number of sites that we have measurements on exceeds the number of samples. Addressing these issues has been a focus of the mediation literature, with authors using penalized regression^34–38^, dimension reduction^39–41^, Bayesian inference^15,42^, and latent variables^43^ to make the outcome model statistically tractable.

We provide a graphical depiction of eleven available methods in Fig. 2, dividing them into **three** different groups. Each method is described in greater detail in the Methods section and up to nine of them are included in the analysis. In the first group, we consider methods that fit the above pair of models explicitly, allowing one to estimate ***α***_***a***_^*T*^***β***_***m***_, the global indirect effect, simply by summing the estimated mediation contributions. These include high-dimensional mediation analysis (HIMA) by Zhang et al. 2016^34^, high-dimensional mediation analysis (HDMA) by Gao et al. 2019^35^, mediation analysis via fixed effect model (MedFix) by Zhang 2019^36^, pathway least absolute shrinkage operator (pathway LASSO) by Zhao and Luo 2022^37^, the Bayesian sparse linear mixed model (BSLMM) by Song et al. 2020^15^, and the Gaussian mixture model (GMM) by Song et al. 2021^42^. In the second group, we consider methods that can estimate ***α***_***a***_^*T*^***β***_***m***_ “directly”; in other words, without needing to fit the original pair of models explicitly. These have the drawback of being unable to identify specific active mediators because they do not provide estimates of the mediation contributions. They include principal component mediation analysis (PCMA) by Huang and Pan 2016^39^, sparse principal component mediation analysis (SPCMA) by Zhao et al. 2020^40^, and high-dimensional linear mediation analysis (HILMA) by Zhou et al. 2021^38^. Last, in the third group, we consider methods that make no attempt to estimate the mediation effects as originally proposed, but instead reconceptualize the mediation framework with newly-defined parameters based on latent variables. This group includes the methods high-dimensional multivariate mediation analysis (HDMM) by Chén et al. 2018^41^ and latent variable mediation analysis (LVMA) by Derkach et al. 2021^43^. Within this comparative structure, we evaluate methods from all three groups, identifying their strengths and weaknesses across a wide range of simulation settings and analysis of DNAm data from MESA.

**Fig. 2.**
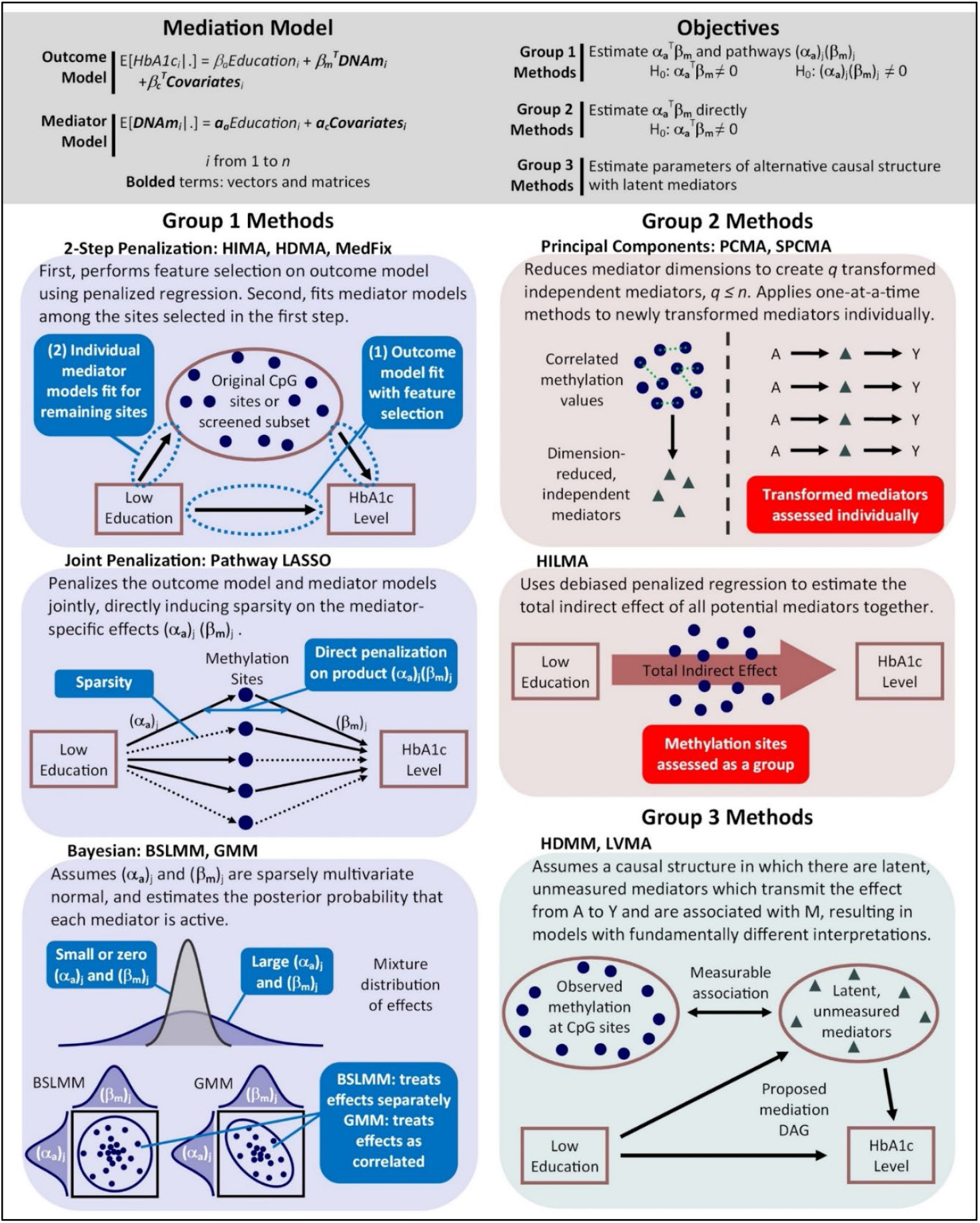
Methods for mediation analysis with high-dimensional DNAm data. Figure describes eleven methods for mediation analysis that can be applied to high-dimensional DNA methylation data, each of which is described in greater detail in the Methods section. Seven of these methods are included in the simulation study and nine in the observed DNAm data analysis with MESA. Group 1 methods fit the outcome model explicitly using penalized regression or Bayesian regresion; Group 2 methods obtain the global mediation effect without fitting the original outcome model explicitly; and Group 3 methods measure mediation through latent variables.

## Results

### Simulation Results

We begin by comparing the performance of the methods using simulations, where we know and can control the true values of the parameters. On simulated data with 2,000 (potential) mediators and either 1,000 or 2,500 observations, we consider (1) a baseline setting, where the mediators are moderately correlated and their signals are sparse; (2) a high-correlation setting, where the correlations between mediators are enhanced compared to (1); and (3) a non-sparse setting, where every mediator has at least some mediation signal but some of the signals are systematically larger. In Settings (1) and (2), 60 random mediators have **(*α***_***a***_)_*j*_ only sampled from a Normal(0,1), 60 have **(*β***_***m***_)_*j*_ only sampled from a Normal(0,1), and 20 have both, with the remaining entries of ***α***_***a***_ and ***β***_***m***_ fixed at zero. In Setting (3), we use a similar scheme, but sample the previously zero **(*α***_***a***_)_*j*_ and **(*b***_***m***_)_*j*_ from a Normal(0,0.2^2^). Our simulations also vary the strength of the signals within each of these settings by changing the proportion of variance that is explained by the associations. We do so by changing PVE_A_, the proportion of variance in each mediator that can be explained by *A*, among those mediators that are affected by *A*; PVE_IE_, the proportion of variance of *Y* that is explained by the total mediation effect; and PVE_DE_, the proportion of variance of *Y* that is explained by the direct effect of *A* on *Y*. Results for varying PVE_IE_ are presented here while results for varying PVE_DE_ and PVE_A_ are included in the supplement (Supplementary Figs 1-4). In addition to the high-dimensional mediation methods, we include a one-at-a-time method^44^ in which the mediators are assessed individually using linear regression. We evaluate the methods by their true positive rate (TPR) for detecting active mediators, their mean squared error (MSE) for estimating the contributions of active and inactive mediators, and their percent relative bias for estimating the global indirect effect. See Methods for more details.

#### True positive rate

Fig. 3 compares the TPR detecting active mediators of the Group 1 methods and the one-at-a-time method. The value shown is the mean TPR over 100 simulated datasets and a 95% empirical confidence interval (CI). On each dataset and for each method, thresholding was used to keep the false discovery rate (FDR) below 10%. For the non-sparse setting, we show the TPR for detecting mediators whose **(*α***_***a***_)_*j*_ and **(*β***_***m***_)_*j*_ were both sampled from Normal(0,1) rather than Normal(0,0.2^2^). We include the Group 1 methods HIMA, HDMA, MedFix, pathway LASSO, and BSLMM. We focus on TPR but not false positive rate (FPR) because the FDR correction was highly conservative, the mean FPR ranging from 0 to 5.1×10^−4^ across all settings and methods.

**Fig. 3.**
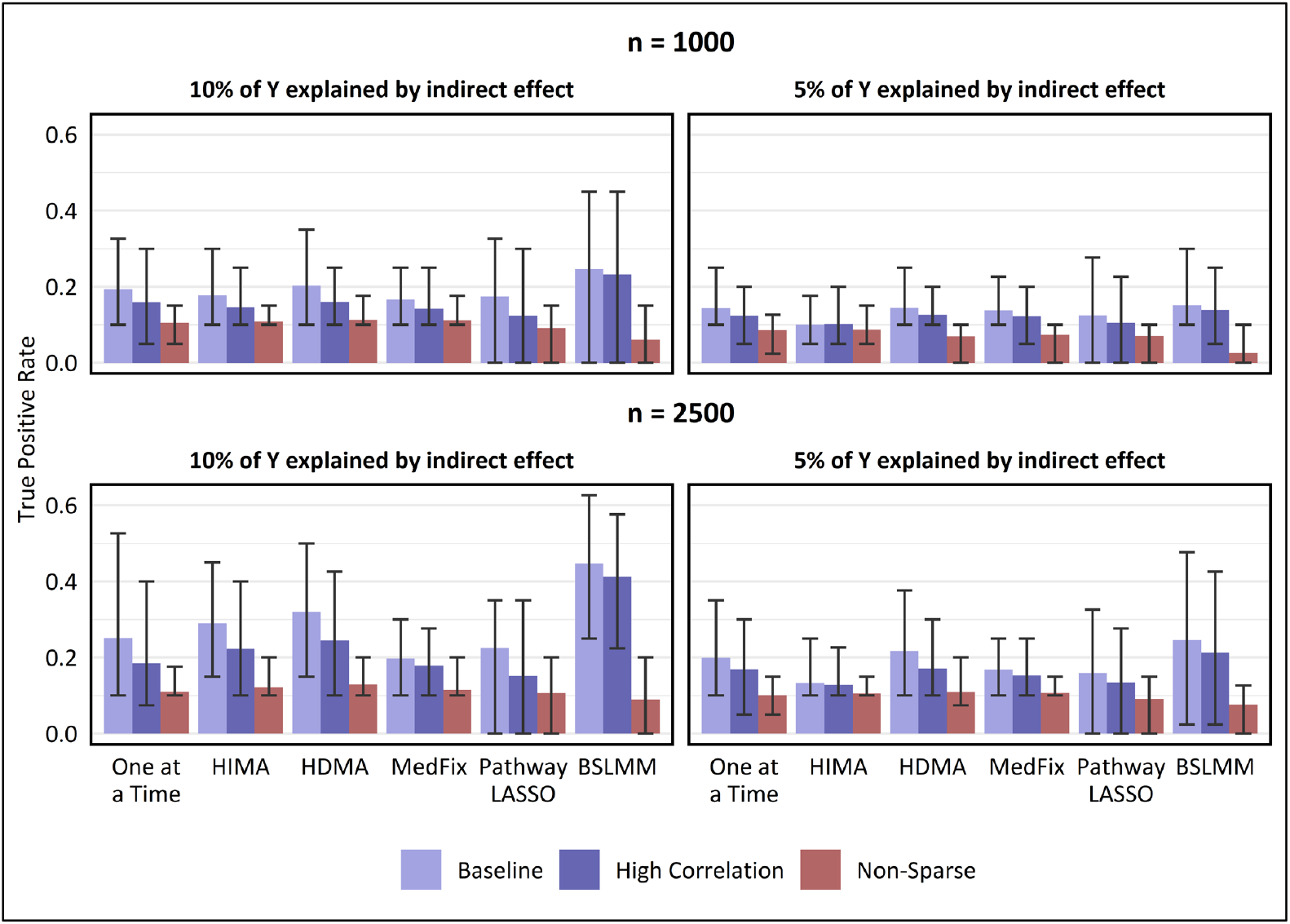
True positive rate for detecting mediation signals at a false discovery rate of 10%. Value shown is the mean TPR across 100 simulated data replicates, with intervals representing the inner 95% range. In the baseline and high-correlation-among-mediators settings, TPR is for distinguishing mediators which contribute to the global mediation effect from those which do not, whereas in the non-sparse setting, TPR is for distinguishing mediators whose contributions were sampled from a high-variance distribution from those whose contributions were sampled from a low-variance distribution. False discovery proportion was capped below 10% by a proper choice of the p-value threshold (one-at-a-time, HIMA, HDMA, MedFix), posterior inclusion probability threshold (BSLMM), or method tuning parameter (pathway LASSO).

For a sample size of 2,500 and a PVE_IE_ of 0.10, the most powerful method in the baseline setting was BSLMM (mean TPR: 0.45; CI: 0.25 - 0.63), whose average TPR was 40% higher than that of the second-best method, HDMA. BLSMM also performed best when PVE_IE_ was 0.05 (mean TPR: 0.25; CI: 0.02 - 0.48), but to a lesser degree, outperforming HDMA by only 13%. BSLMM remained the best method, and HDMA the second best, no matter the signal strength or the degree of correlations, but performed poorly when the signals were non-sparse. In the setting with 1,000 observations, PVE_IE_ set to 0.05, and non-sparse signals, the best-performing method was HIMA (mean TPR: 0.09; CI: 0.05 - 0.10), its average TPR 3.3 times higher than that of BSLMM, which performed worst.

#### Estimation of contributions of active mediators

Next, we assess the MSE of the methods for estimating mediation contributions, relative to the one-at-a-time approach. In Fig. 4, we show the relative MSE (rMSE) for estimating mediation contributions among the mediators that were either active (in the baseline and high-correlation settings) or had (***α***_***a***_)_*j*_ or (***β***_***m***_)_*j*_ sampled from the larger-variance distribution (in the non-sparse setting). In the baseline setting with 2,500 observations, the best-performing method when the mediation signal was strong was BSLMM, whose mean rMSE of 0.59 (CI: 0.13 - 1.51) was 24% lower than that of HDMA, the second-best method. However, when the PVE_IE_ was reduced to 0.05 or the sample size reduced to 1,000, the best-performing method was either HDMA or MedFix, with MedFix (mean rMSE: 0.79; CI: 0.31 - 1.53) performing 61% better than BSLMM after reducing both. Similar trends were observed for the high-correlation and non-sparse settings.

**Fig. 4.**
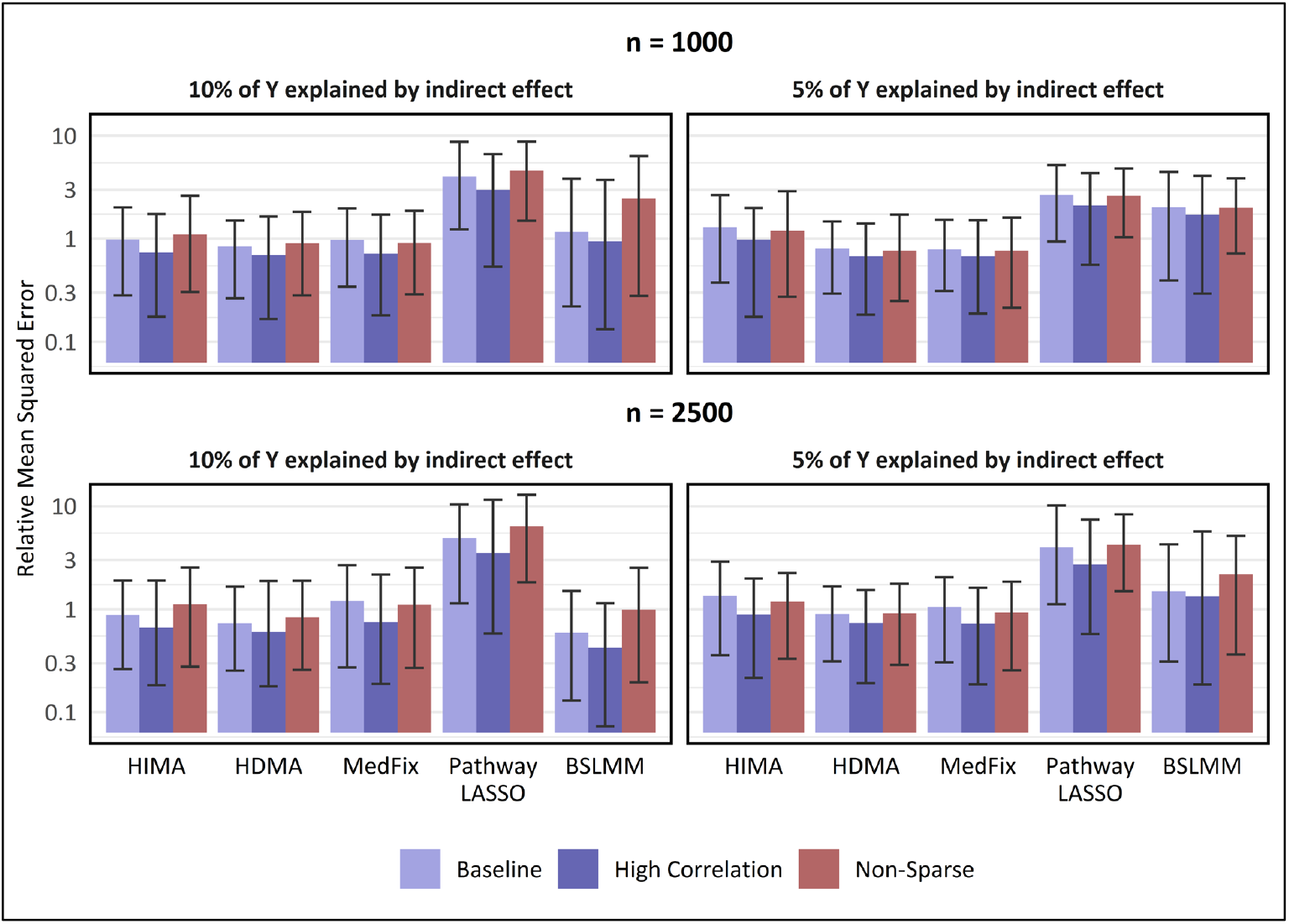
MSE in estimating mediation contributions of active mediators, relative to one-at-a-time method. Y-axis is on a log_10_ scale. Value shown is the mean of the relative mean-squared error for estimating mediation contributions among active mediators (relative to the one-at-a-time approach) across 100 simulated data replicates, with intervals representing the inner 95% range. For baseline and high-correlation-between-mediators settings, active mediators which contribute to the global mediation effect, whereas in the non-sparse setting, active mediators are those whose contributions were sampled from a distribution with large variance instead of small.

#### Estimation of contributions of inactive mediators

Figure 5 shows the rMSE among the mediators that either were not active (in the baseline and high-correlation settings) or had **(*α***_***a***_)_*j*_ or **(*β***_***m***_)_*j*_ sampled from the smaller-variance distribution (in the non-sparse setting). We exclude pathway LASSO from Fig. 4 because for the baseline and high-correlation settings it had rMSEs of exactly zero. The reason for this is that pathway LASSO tended to be highly conservative and successfully assigned inactive mediators to have no effect. As for the other methods, in the baseline setting with 2,500 samples, MedFix performed the best when PVE_IE_ was 0.10, with a mean rMSE of 1.8×10^−3^ (CI: 1.9×10^−4^ - 6.4×10^−3^), which was 46% lower than the mean rMSE for the second-best method, HIMA. In contrast, HIMA was the best-performing method when signal was weakened to a PVE_IE_ of 0.05, attaining a mean rMSE of 2.8×10^−4^ (CI: 0.0 - 1.3×10^−3^), which was 94% lower than that of the second-best, MedFix. Results were similar when the correlations between mediators were heightened and when the sample size was reduced. In the settings where mediation signals were non-sparse, the best-performing method was always HIMA, which had a mean rMSE of 3.7×10^−2^ (CI: 1.1×10^−2^ - 6.5×10^−2^) when PVE_IE_ was 0.10 and there were 2,500 observations, 2% lower than that of MedFix.

**Fig. 5.**
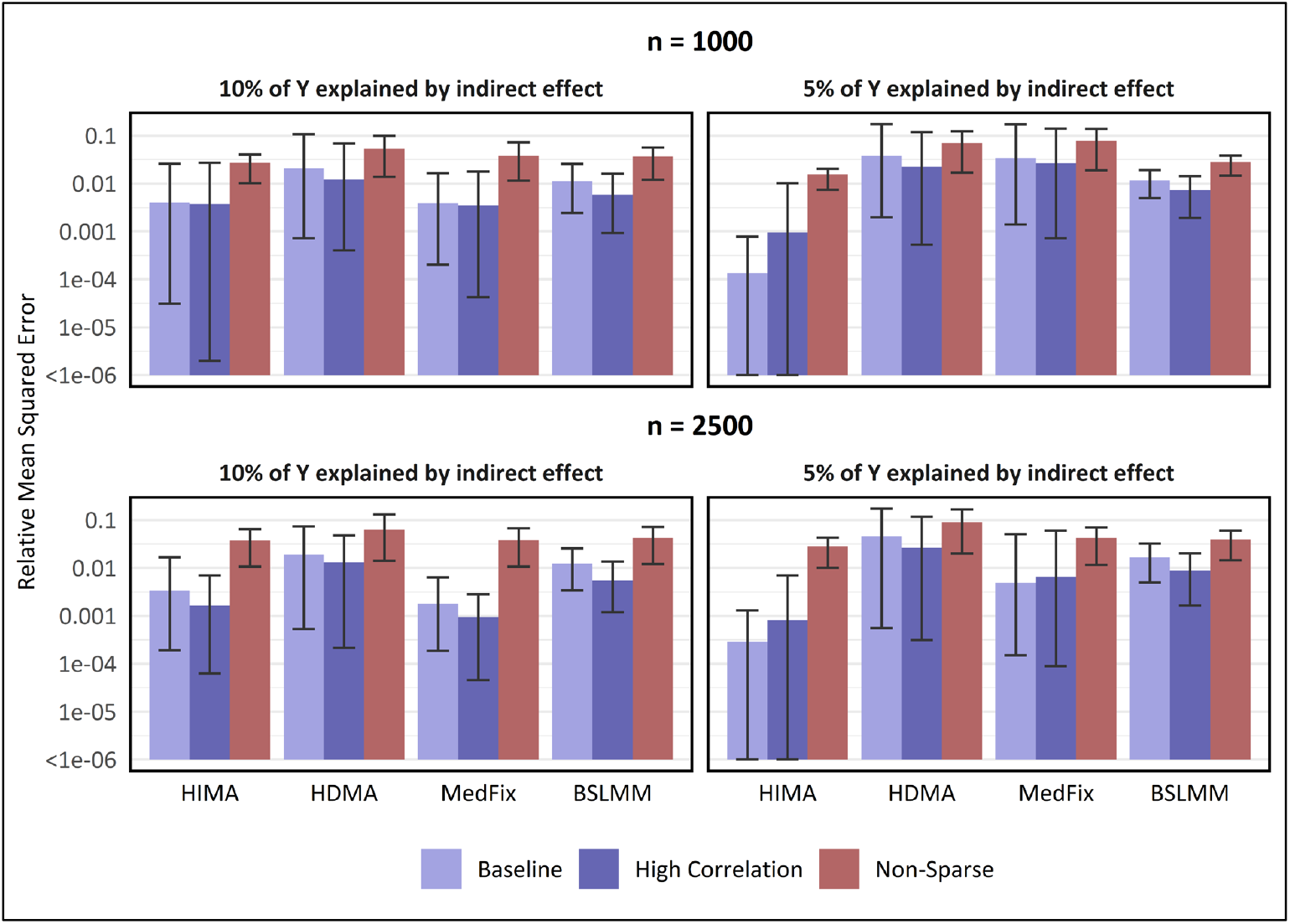
MSE in estimating mediation contributions of inactive mediators, relative to one-at-a-time method. Y-axis is on a log_10_ scale. Value shown is the mean of the relative mean-squared error for estimating mediation contributions among inactive mediators (relative to the one-at-a-time approach) across 100 simulated data replicates, with intervals representing the inner 95% range. For baseline and high-correlation-between-mediators settings, inactive mediators are those which do not contribute to the global mediation effect, whereas in the non-sparse setting, inactive mediators are those whose contributions were sampled from a distribution with small variance instead of large.

#### Estimation of global indirect effect

Lastly in Fig. 6, we show the percent relative bias for estimating ***α***_***a***_^**T**^***β***_***m***_, the global indirect effect. We use the same methods as in Figures 3 to 5 along with the Group 2 methods PCMA and HILMA, which obtain an estimate of the global indirect effect without needing to directly fit the original mediation model. (The Group 2 method SPCMA is excluded for computational reasons.) In the baseline setting with 2,500 samples, the best performer when PVE_IE_ was 0.10 was HILMA, whose mean relative bias of 9% (CI: 0.6% - 20.8%) was 40% lower than that of HDMA, the second-best. Next, when the PVE was reduced to 0.05, the best-performing method was MedFix (mean relative bias: 20.5%; CI: 1.0% - 43.8%), which outperformed HILMA by only 7%. We observed similar results for a sample size of 1,000 and high-correlations. In the non-sparse settings, where the biases tended to be much higher, the best performing methods were either PCMA or HDMA.

**Fig. 6.**
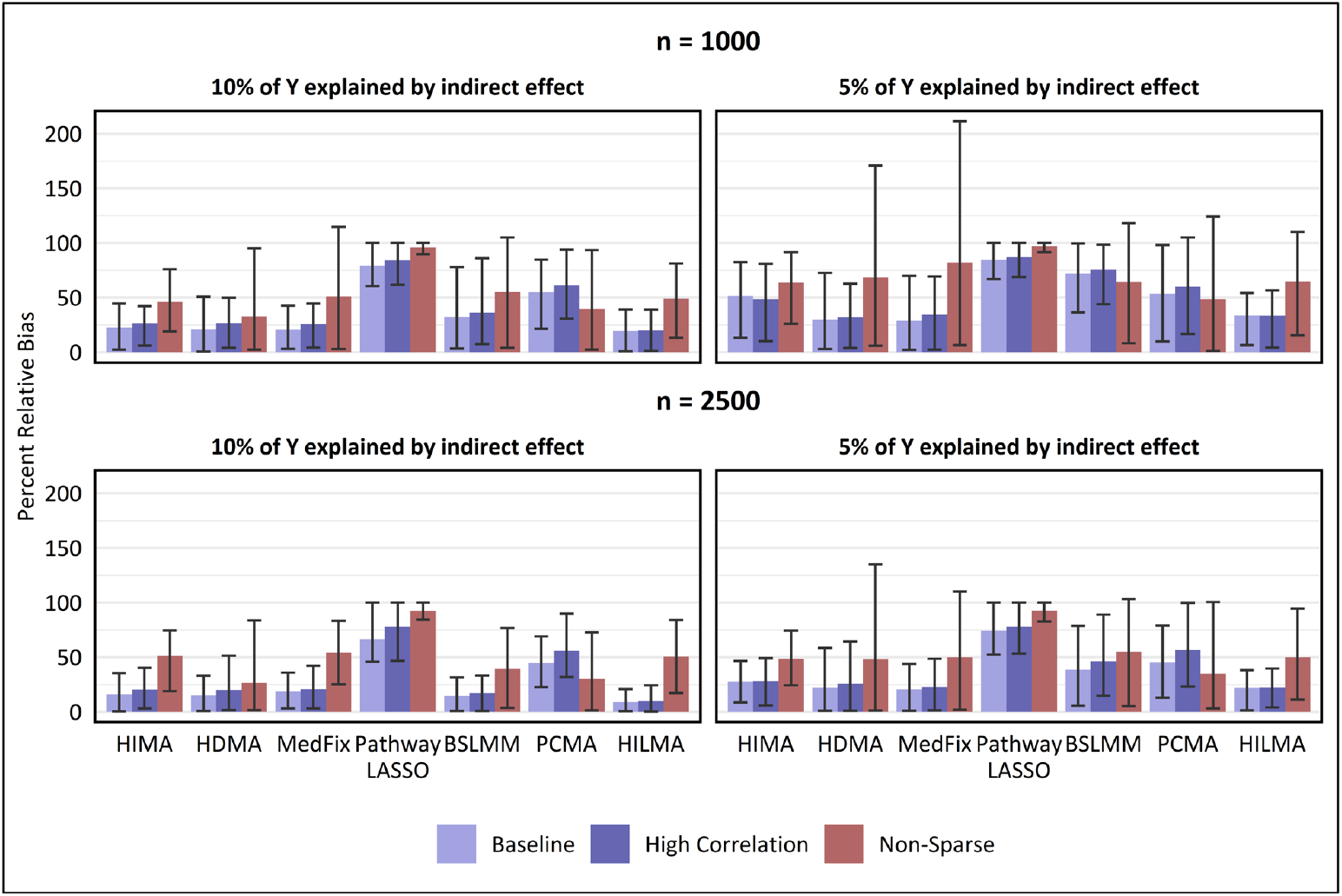
Percent relative bias in estimated global indirect effect. Value shown is the mean of the percentage relative bias in estimating the global mediation effect across 100 simulated data replicates, with intervals representing the inner 95% range.

#### Scalability

We evaluated the scalability of the methods by running them 30 times on a common computing platform, and recording their run time (Table 1). This was done in both a small data setting (*n* = 100, *p* = 200) and a big data setting (*n* = 1,000, *p* = 1,000). On the larger dataset, the methods MedFix, HDMA, and PCMA posed insignificant computational burden; whereas BSLMM took an average of 40.1 minutes per run (assuming 30,000 posterior samples), HILMA an average of 40.9 minutes per run, pathway LASSO an average of 192.6 minutes per run, and SPCMA an average of 842.5 minutes per run (assuming 100 principal components). Run times were substantially lower in the smaller dataset, the slowest method, pathway LASSO, only taking an average of 18.71 minutes. The memory consumption of the methods is included in Supplementary Table 1.

**Table 1.** Computation time comparison for high-dimensional mediation analysis methods.

| Method | $n = 100, p = 200$ | | $n = 1,000, p = 2,000$ | |
| --- | --- | --- | --- | --- |
|  | Mean | Interquartile Range | Mean | Interquartile Range |
| BSLMM | 39.17s | (38.84s - 39.54s) | 40.14m | (39.74m - 40.34m) |
| HDMA | 1.40s | (1.37s - 1.40s) | 29.76s | (29.55s - 29.92s) |
| HDMM | 24.85s | (24.80s - 24.89s) | 12.36m | (12.33m - 12.37m) |
| HILMA | 24.42s | (24.13s - 24.63s) | 40.85m | (38.22m - 40.65m) |
| HIMA | 0.25s | (0.25s - 0.25s) | 3.55s | (3.47s - 3.62s) |
| MEDFIX | 0.61s | (0.60s - 0.61s) | 7.33s | (7.22s - 7.42s) |
| PCMA | 2.77s | (2.74s - 2.79s) | 58.97s | (58.08s - 59.35s) |
| PLASSO | 18.71m | (18.19m - 19.23m) | 192.62m | (188.10m - 195.83m) |
| SPCMA | 16.05m | (15.94m - 16.04m) | 842.54m | (827.26m - 855.21m) |

Methods were run 30 times each on a single core of an Intel(R) Xeon(R) Gold 6242R CPU @ 3.10GHz processor.

### DNAm data analysis results from MESA

For our real data analysis, we applied the methods on a dataset with high-dimensional epigenetic mediators. Our exposure of interest was low SES—measured by educational attainment below a four-year degree—while our outcome variable was HbA1c level and our potential mediators were DNAm measurements at 402,339 CpG sites. Since the methods are incapable of handling so many CpG sites at once, we reduced our scope to only include the 2,000 sites with the strongest association with low SES. This was based on a linear mixed-model adjusting for age, sex, race, and the estimated proportions of residual non-monocytes as fixed effects and methylation chip and position as random effects. Our final dataset contained these 2,000 CpG sites and 963 samples. HbA1c, DNAm, and all other continuous variables were standardized prior to analysis.

#### Identification of noteworthy CpG sites

We identified CpG sites that potentially mediated the relationship between low SES and HbA1c using the Group 1 methods HIMA, HDMA, MedFix, pathway LASSO, and BSLMM. In HIMA, HDMA, MedFix, and pathway LASSO, which involve feature selection, we describe a CpG site to be “active” if its estimated mediation contribution is not zero; whereas in BSLMM, we do so if the estimated posterior inclusion probability is not zero (see Methods). We also included a one-at-a-time method in which the CpG sites were assessed individually with linear mixed models, identifying active mediators with the joint significance test^44^. Out of 2,000 CpG sites, HIMA found 3 sites to be noteworthy, HDMA found 11, MedFix found 3, pathway LASSO found 141, and BSLMM found 3, amounting to 144 unique CpG sites in total. The one-at-a-time method identified zero CpG sites as noteworthy at an FDR threshold of 10%. Eleven CpG sites were identified as noteworthy by at least two of the methods (Table 2). Among these 11, the estimated mediation contributions were similar across methods in direction and size except for BSLMM, for which the estimates were an order of magnitude smaller than the others but in the same direction.

**Table 2.** Estimated contributions of noteworthy CpG sites on the mediation pathway between low education and HbA1c.

| CpG Name | Chromosome | Nearby Gene(s) | USCS RefGene Group | Univariate (0 sites identified) | HIMA (3 sites identified) | HDMA (11 sites identified) | MedFix (3 sites identified) | Pathway LASSO (141 sites identified) | BSLMM (3 sites identified) |
| --- | --- | --- | --- | --- | --- | --- | --- | --- | --- |
| cg10508317 | 17 | *SOCS3* | Body | $3.48 \times 10^{-2}$ | $1.59 \times 10^{-2*}$ | $3.56 \times 10^{-2*}$ | $2.90 \times 10^{-2*}$ | $2.35 \times 10^{-2*}$ | $0.25 \times 10^{-2}$ |
| cg01288337 | 14 | *RIN3* | Body | $3.35 \times 10^{-2}$ | $1.47 \times 10^{-2*}$ | $2.82 \times 10^{-2*}$ | $2.70 \times 10^{-2*}$ | $4.43 \times 10^{-2*}$ | $0.21 \times 10^{-2}$ |
| cg10244976 | 16 | *LMF1* | Body | $3.00 \times 10^{-2}$ | 0 | $2.78 \times 10^{-2*}$ | 0 | $2.23 \times 10^{-2*}$ | $0.19 \times 10^{-2}$ |
| cg07516252 | 14 | *REC8* | TSS200 | $2.72 \times 10^{-2}$ | 0 | $2.24 \times 10^{-2*}$ | 0 | $2.26 \times 10^{-2*}$ | $0.26 \times 10^{-2}$ |
| cg07571519 | 10 | *C10orf105*; *CDH23* | 3'UTR; Body | $2.53 \times 10^{-2}$ | $0.33 \times 10^{-2*}$ | $3.67 \times 10^{-2*}$ | $1.47 \times 10^{-2*}$ | $2.81 \times 10^{-2*}$ | $0.21 \times 10^{-2}$ |
| cg23079012 | 2 | *LINC00299* | Body | $2.27 \times 10^{-2}$ | 0 | $1.99 \times 10^{-2*}$ | 0 | $1.98 \times 10^{-2*}$ | $0.29 \times 10^{-2}$ |
| cg01587454 | 8 | *DCAF4L2* | 1stExon | $1.77 \times 10^{-2}$ | 0 | $2.10 \times 10^{-2*}$ | 0 | $1.99 \times 10^{-2*}$ | $0.38 \times 10^{-2}$ |
| cg27527503 | 4 | *HADH* | TSS1500 | $1.75 \times 10^{-2}$ | 0 | $1.86 \times 10^{-2*}$ | 0 | $1.27 \times 10^{-2*}$ | $0.23 \times 10^{-2}$ |
| cg25891647 | 11 | *GRAMD1B* | Body | $-1.27 \times 10^{-2}$ | 0 | $-3.42 \times 10^{-2*}$ | 0 | $-3.02 \times 10^{-2*}$ | $-0.33 \times 10^{-2}$ |
| cg08473752 | 17 | *NLK* | Body | $-0.70 \times 10^{-2}$ | 0 | $-2.34 \times 10^{-2*}$ | 0 | $-2.32 \times 10^{-2*}$ | $-0.22 \times 10^{-2}$ |
| cg12644059 | 15 | *BLM* | N/A<sup>1</sup> | $-0.03 \times 10^{-2}$ | 0 | $-2.31 \times 10^{-2*}$ | 0 | $-1.84 \times 10^{-2*}$ | $-0.22 \times 10^{-2}$ |

\*Selected as noteworthy by given method
<sup>1</sup>CpG site cg12644059 is 3.240kb from the final base pair of the BLM gene
Table includes all CpG sites that were selected as having a noteworthy mediation contribution by at least two of the implemented methods out of 2,000 CpG sites in total. Criteria for CpG identification varied by method. All estimates are adjusted for age, sex, race, and the estimated proportions of residual non-monocytes as fixed effects, along with methylation chip and position as random effects to address potential batch effects. Note that for HIMA, HDMA, MedFix, and pathway LASSO, which fit high-dimensional regression models, we used additional pre-screening to reduce the number of mediators in advance to only $n/\log(n) \approx 141$ CpG sites, which is the approach recommended by the HIMA and HDMA authors and helps with statistical and computational efficiency (see Methods). Pathway LASSO selected all of these 141.

Some of these CpG sites were on or nearby genes that are potentially related HbA1c. Site cg10508317 is in the body of the *SOCS3* gene, for which a rich body of literature has established links between overexpression and insulin resistance^45^. The same site has also been identified in MESA as a mediator between adult SES and BMI^46^ and adult SES and HbA1c^31^ based on previous one-at-a-time analyses. Site cg01288337, in the body of the *RIN3* gene, has been identified in MESA as a potential mediator between adult SES and HbA1c based on one-at-a-time analysis as well^31^. The *RIN3* gene itself is proximal to the *SLC24A4* gene, both of which have been linked to brain glucose metabolism in human population studies^47^. In addition, site cg27527503 is in the promoter region of the *HADH* gene, which is differentially expressed with respect to diabetes status^48^ and is a primary driver of hyperinsulinism^49^ and hyperinsulinaemic hypoglycemia (low blood sugar due to excess insulin)^50^. A Venn diagram of genes identified by the methods is included in Supplementary Fig. 5, and results for every noteworthy CpG site are listed in Supplement File 1.

#### Global mediation through DNAm

Next, we estimated the direct effect of low education on HbA1c, the global indirect effect of low education on HbA1c through DNAm, and the total effect of low education on HbA1c using the Group 1 methods HIMA, HDMA, MedFix, pathway LASSO, and BSLMM, as well as the Group 2 methods PCMA, SPCMA, and HILMA (Table 3). Results across methods varied considerably, with the estimated global indirect effect ranging from 0.03 in HILMA to 0.17 in SPCMA. The estimated total effect ranged from 0.02 (HILMA) to 0.198 (HIMA, HDMA, and MedFix). While HILMA appeared to be an outlier, some of the other methods were consistent, with HDMA, BSLMM, P-LASSO, PCMA, and SPCMA all estimating the global indirect effect to be close to 0.15. The variability in the estimated indirect effect and estimated total effect led to variability in the proportion mediated as well, from 17.1% in HIMA to 100% in HILMA.

**Table 3.** Estimated effects in the mediation mechanism from low education to DNAm to HbA1c.

| Method | Estimated Global indirect Effect | Estimated Direct Effect | Estimated Total Effect | Estimated Proportion Mediated |
| --- | --- | --- | --- | --- |
| HIMA | 0.03 | 0.16 | 0.20 | 0.17 |
| HDMA | 0.13 | 0.07 | 0.20 | 0.65 |
| MedFix | 0.07 | 0.13 | 0.20 | 0.36 |
| BSLMM | 0.14 | 0.05 | 0.18 | 1.00 |
| Pathway LASSO | 0.13 | 0.05 | 0.18 | 0.74 |
| PCMA | 0.15 | 0.02 | 0.17 | 0.91 |
| SPCMA | 0.17 | 0.00 | 0.17 | 1.00 |
| HILMA | 0.03 | 0.00 | 0.03 | 1.00 |

All estimates are adjusted for age, sex, race, and the estimated proportions of residual non-monocytes as fixed effects, along with methylation chip and position as random effects to address potential batch effects. We provide only point estimates, not interval estimates, because some of the methods are either not capable of producing interval estimates or do not provide the code for producing them in their software. For HIMA, HDMA, and MedFix, which as coded do not directly provide estimates of the direct effect, we first estimate the total effect by fitting the outcome model with the CpG sites omitted, then estimate the direct effect by subtracting the indirect effect from the total effect. Note also that, for HIMA, HDMA, MedFix, and pathway LASSO, we used additional screening to reduce the number of mediators in advance for the sake of statistical and computational efficiency, so only $n/\log(n) \approx 141$ CpG sites were seen by the multivariate model rather than 2,000 (this approach is recommended by the HIMA and HDMA authors).

#### Additional Findings

In addition to estimating the global indirect effect, method SPCMA is also able to identify potentially-mediating CpG sites in groups. It does so by linearly combining the mediators using sparse principal component-defined weights, then evaluating the resulting principal components as mediators themselves^40^. However, out of 100 computed principal components, only three of them had significant mediation contributions after 10% FDR correction, the first representing a linear combination of 762 CpG sites, the second a combination of 782 sites, and the third a combination of 797 sites. Since the transformed mediators are functions of so many CpG sites at once, one cannot make claims about which particular CpG sites are active mediators, but the method still provides insight to whether there is statistical mediation at all.

We finish our analysis by deploying HDMM, a method from Group 3. Unlike the methods in Groups 1 and 2, HDMM cannot be used to estimate the global indirect effect from the proposed mediation structure, nor to estimate the mediation contributions of specific CpG sites. Rather, HDMM uses a likelihood-based approach to compute “directions of mediation”, which are weights that can be used to linearly combine the observed mediators into unobserved, latent mediators that replace the observed mediators in the mediation models (similar to PCMA). The estimated effect of the first latent mediator on average HbA1c was 0.13, the estimated total effect 0.71, and the proportion mediated 0.715. The three CpG sites with the largest directions of mediation were cg01288337 (0.36) on the *RIN3* gene, cg16162970 (−0.22) near the *PACS2* gene, and cg25891647 (−0.21) on the *GRAMD1B* gene; the first and last of which were among the 11 CpG sites identified by other methods in Table 2. Although the size and direction of these estimates are not interpretable, they offer evidence that these CpG sites are potentially involved in mediation.

## Discussion

In this study, we reviewed and evaluated statistical methods for performing mediation analysis with high-dimensional DNAm data, so that researchers in epigenetics have the information they need to choose the most appropriate method for their data sample, subject matter, and research objectives. In extensive simulations, we found that the most powerful method for identifying active mediators was generally BSLMM, with HDMA close behind; though the former performed poorly in settings where the mediation signals were non-sparse. No method was uniformly better than the others at estimating the mediation contributions, though pathway LASSO was always the weakest. For estimating the global indirect effect, the best-performing method was HILMA in sparse mediation settings and PCMA or HDMA in non-sparse settings. Our scalability comparison revealed that HIMA, HDMA, MedFix, and PCMA were easily scalable to large datasets (e.g., *n* = 1,000 and *p* = 2,000), whereas SPCMA and pathway LASSO were extremely computationally costly.

On DNAm data from MESA, 11 CpG sites were selected by at least two of the methods as mediators between low SES and HbA1c level. Of the many genes related to these sites, *SOCS3, RIN3*, and *HADH* have the strongest potential biological connections to HbA1c^45,47,48,50–52^, which contributes to the already rich literature on DNAm as a mediator between the exposome and health outcomes.

Moreover, the methods generally produced similar estimates of the mediation contributions, with the exception of BSLMM. It is possible that since BSLMM is non-sparse, the estimated mediation contributions end up severely shrunken compared to the methods which directly select features.

Estimates of the global indirect effect were highly variable. Part of this can be explained by the fact that HDMA, MedFix, HIMA, and pathway LASSO are sparse models that can set mediation contributions to be exactly zero, resulting in a rigid and unstable estimation of the global indirect effect. The method HILMA, which is built specifically for estimating the global indirect effect and direct effect, produced estimates that were sharply different than the other methods, possibly because our simulations indicated that it struggled in non-sparse mediation settings.

In practice, the optimal method for mediation analysis with high-dimensional mediators will depend both on the data and the objective. If the goal is to identify specific CpG sites that are involved in mediation, one preferred method may be HDMA, which performed well at detecting active mediators in our simulations and was not overly conservative when applied to the observed data. If one’s focus is the global indirect effect, our simulations suggested that the optimal method is HILMA; but considering the variability we observed in our DNAm analysis, it may be worthwhile to apply BSLMM and HDMA as well to ensure the results are robust. If the results of multiple methods disagree substantially, it may be difficult to say with confidence which is closest to the truth, and the estimates should be interpreted with caution. Next, if there is interest in latent, unmeasured mediators, either HDMM or LVMA is worth attempting, though HDMM is computationally simpler. A detailed decision tree for selecting the optimal method is included in Fig. 7.

**Fig. 7.**
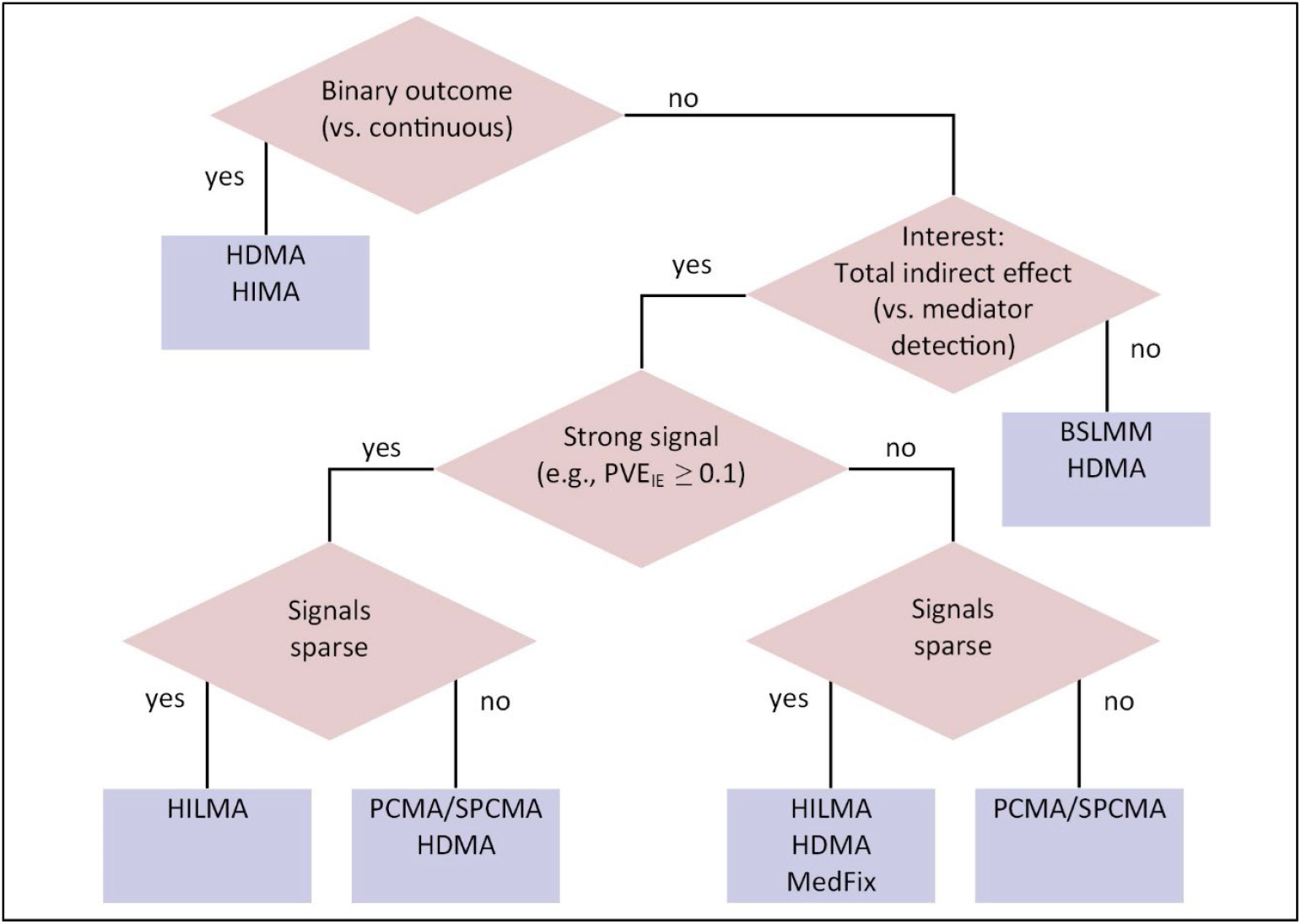
Decision tree for selecting a high-dimensional mediation analysis.

Some strengths of our study include its broad coverage of the available methods, the breadth of its simulation settings, and the comprehensive set of evaluation criteria. Our analysis of real DNAm data is especially essential because it elucidates the potential limitations of using these methods in practice, as it is impossible to incorporate the full complexity of real data sources into contrived simulation settings. However, our study also has weaknesses. First, since DNAm measurements and HbA1c data were collected concurrently, and represent only single time points, we cannot interpret the parameters we have estimated as causal effects. Nor can we interpret the mediation contributions estimated in Table (2) as causal, since DNAm was correlated across CpG sites and we have made no assumptions about their causal ordering. Moreover, although it would be optimal to address our research question longitudinally, with measurements at multiple time points, there is a dearth of mediation analysis methods which can handle that type of data, and longitudinal mediation analysis with high-dimensional mediators should be a focus of future methodological development. Second, we limited our analysis to the situation that *Y* and ***M*** are continuous, that ***M*** and *A* do not interact, and that only one *A* is of interest. However, we note that the methods HIMA and HDMA can also be applied to identify active mediators when *Y* is binary, while PCMA can be applied to infer the global indirect effect when there is *A*-***M*** interaction in the outcome model. MedFix, along with the simultaneously-proposed MedMix (mediation analysis with mixed effect model by Zhang (2021)) can be applied when both the exposures and mediators are high-dimensional, while Huang and Vanderweele (2014) proposed a variance component test of the global indirect effect when only *A* is high-dimensional^53^. As the landscape of methods for high-dimensional mediation analysis continues to expand, future review studies should consider exploring additional mediation settings (in presence of non-linearity, interaction) for which statistical methods are continuing to become available.

## Methods

### Mediation Model with Multiple Mediators

Let ***M*** be a set of *p* variables, *M*^(1)^, *M*^(2)^, to *M*^(*p*)^, each a potential mediator in the causal pathway between *A* and *Y*. We assume that the ordering of the potential mediators is arbitrary and that *Y* is continuous. Given a dataset of *n* individuals, with *A*_*i*_, *Y*_*i*_, ***M***_*i*_, and *q* covariates ***C***_*i*_ measured for each subject *i*, we can evaluate the mediating role of ***M*** with the models

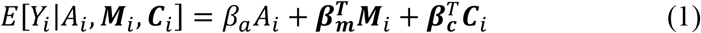

and

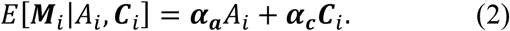

We refer to these as the outcome and mediator models. Bolded terms distinguish vectors from scalars. Under certain assumptions, the parameters of this model can be used to derive causal effects of interest: Namely, in addition to the baseline assumption of temporality, we assume (1) that there is no unmeasured confounding in the exposure-outcome association after conditioning on ***C***, (2) that there is no unmeasured confounding in the mediator-outcome associations after adjusting for the exposure and ***C***, (3) that there is no unmeasured confounding of the exposure-mediator associations after conditioning on ***C***, and (4) that the measured confounders of the mediator-outcome associations are not caused by the exposure (which would make those confounders mediators themselves). In these circumstances only can *β*_*a*_ be interpreted as the natural direct effect of *A* on *Y*, ***α***_***a***_^*T*^***β***_***m***_ the natural indirect effect of *A* on *Y* through ***M***, and *β*_*a*_ *+* ***α***_***a***_^*T*^***β***_***m***_ the total effect of *A* on *Y*^33^. We say a mediator *M*^(*j*)^ is *active* if (***α***_***a***_)_*j*_(***β***_***m***_)_*j*_ is not zero, since it contributes mathematically to the indirect effect, but this contribution itself cannot be formally interpreted causally unless the mediators are independent conditional on *A* and ***C***. Extensions of this framework cover cases when *Y* is binary, when ***M*** is binary, or when the outcome model requires an interaction effect between ***M*** and *A*^33^.

A summary of the methods that can evaluate ***M*** as a mediator is provided in Table 4, using the above pair of models as a frame of reference. We describe each of the methods in greater detail in the following three sections.

**Table 4.** Methods Summary.

| Name and Author | Estimation of global indirect effect | Estimation of mediation contributions | Mediator identification | *Y* Data Type | Summary |
| --- | --- | --- | --- | --- | --- |
| Group 1 Methods |  |  |  |  |  |
| HIMA; Zhang, 2016 | Point estimation | Point estimation | Yes | Continuous or binary | Fits the outcome model with the minimax concave penalty. Requires subsequent fitting of ordinary least squares regression to test the statistical significance the mediation contributions. |
| HDMA; Gao, 2019 | Point estimation | Point, interval estimation | Yes | Continuous or binary | Fits the outcome model with the de-sparsified LASSO penalty. |
| MedFix; Zhang, 2021 | Point estimation | Point, interval estimation | Yes | Continuous | Fits the outcome model with the adaptive LASSO penalty. Can also be applied when the exposure is high-dimensional in addition to the mediators. |
| Pathway LASSO Zhao and Luo, 2022 | Point estimation | Point estimation | Yes | Continuous | Fits the outcome model and mediator models with a jointly penalized likelihood, directly applying shrinkage to the mediation contributions $(\alpha_a)_j(\beta_m)_j$. |
| BSLMM; Song, 2020 | Bayesian point, interval estimation | Bayesian interval estimation | Yes | Continuous | Bayesian mixed-model in which the mediator-outcome associations $(\beta_m)_j$ and the exposure-mediator associations $(\alpha_a)_j$ are assumed to independently follow sparse normal distributions. |
| GMM; Song, 2021 | Bayesian point, interval estimation | Bayesian interval estimation | Yes | Continuous | Bayesian mixed-model in which the mediator-outcome associations $(\beta_m)_j$ and the exposure-mediator associations $(\alpha_a)_j$ are assumed to jointly follow a sparse multivariate normal distribution. |
| Group 3 Methods |  |  |  |  |  |
| PCMA; Huang and Pan, 2016 | Point, interval estimation | No | No | Continuous or binary | Applies principal component analysis on the mediator model residuals, transforming the mediators so they are independent. Can be applied when there is $A$-$M$ interaction in the outcome model. |
| SPCMA; Zhao, 2019 | Point, interval estimation | No | Identifies whether subsets of the mediators are jointly active | Continuous | Similar to PCMA but applies sparse PCA, resulting in transformed mediators that are more interpretable. |
| HILMA; Zhou, 2020 | Point, interval estimation | No | No | Continuous | Uses a debiased penalized regression approach to directly estimate the global indirect effect $\alpha_a^T \beta_m$. Can be applied for multiple exposures simultaneously. |
| Group 3 Methods |  |  |  |  |  |
| HDMM; Chen, 2018 | No | No | Nonspecifically identifies groups of active mediators | Continuous | Estimates “directions of mediation” by which the observed mediators can be linearly combined to form latent mediators. The latent mediators replace the true mediators in the analysis. |
| LVMA; Derkach, 2019 | No | No | Identifies inputted mediators associated with latent mediators | Continuous or binary | Reformulates the causal structure of the mediation problem. Assumes that $M$ itself is not responsible for mediation, but rather that the effect of $A$ on $Y$ is mediated by latent, unmeasured factors, $F$, which also cause changes in $M$. |

### Group 1 Methods

This group of methods can estimate both the global indirect effect ***α***_***a***_^*T*^***β***_***m***_ and the mediator-specific contributions **(*α***_***a***_)_*j*_**(*β***_***m***_)_*j*_, *j* from 1 to *p*.

#### HIMA

High-dimensional mediation analysis (HIMA), proposed by Zhang et al. (2016), is a penalized regression approach with two main steps: First, the outcome model is fitted with a minimax concave penalty^54^, performing feature selection on the mediators by setting some of them to have no effect on *Y*^34^. Then, among the remaining mediators, they fit the mediator models individually using ordinary regression. The authors test the significance of **(*α***_***a***_)_*j*_**(*β***_***m***_)_*j*_ by applying Bonferroni correction to the maximum of the **(*β***_***m***_)_*j*_ and **(*α***_***a***_)_*j*_ p-values. To obtain p-values for the **(*β***_***m***_)_*j*_ estimates, the authors re-fit the reduced outcome model by ordinary least squares, which statistically may be overconfident. The authors also recommend an initial screening step to reduce the number of mediators at the start, as the outcome model will still be unstable if *p* is extremely large compared to *n*.

#### HDMA

High-dimensional mediation analysis (HDMA), proposed by Gao et al. (2019), is the same as HIMA except for its penalty function, replacing the minimax concave penalty with the recently-proposed de-sparsified LASSO^35,55^. The advantage of this penalty is that the resulting estimates of ***β***_***m***_ are asymptotically normal, so one can test their statistical significance without needing to subsequently apply ordinary least squares. HDMA is also less biased than HIMA when the mediators are highly-correlated.

#### MedFix

Mediation analysis via fixed effect model (MedFix) is another extension of HIMA, proposed by Zhang (2021)^36^. MedFix was originally proposed for a setting where there are not only multiple mediators, but also multiple exposures, which it handles by applying adaptive LASSO to both the outcome model and the mediator models. If there is only one exposure, feature selection in the mediator models is not necessary, and applying MedFix is analogous to applying HDMA except with adaptive LASSO instead of debiased LASSO.

#### Pathway LASSO

Pathway LASSO is another penalized regression approach, proposed by Zhao and Luo (2022)^37^. Whereas HIMA, HDMA, and MedFix use a two-step design—the outcome model and mediator models fitted separately—this method fits the models all together, with a jointly penalized likelihood. The penalty not only applies shrinkage to the mediator-outcome associations, like the other methods, but also to the exposure-mediator associations and the mediation contributions.

#### BSLMM

The Bayesian sparse linear mixed model (BSLMM) is a Bayesian approach proposed by Song et al. (2020)^15^. The model assumes ***α***_***a***_ and ***β***_***m***_ are random vectors, both independently following mixtures of normal distributions. Most of the effects are presumed to be small, owing to a normal distribution with mean zero and small variance, while the others are allowed to be larger, resulting from a normal distribution with higher variance. We estimate the effects with their posterior mean, and we distinguish active mediators from inactive with their posterior inclusion probability of belonging to the distribution with higher variance.

#### GMM

The Gaussian mixed model (GMM), proposed by Song et al. (2021), is an extension of BSLMM in which the **(*α***_***a***_)_*j*_, **(*β***_***m***_)_*j*_ pairs are treated as correlated, following a mixture of multivariate normal distributions instead of two independent normal distributions^42^. Thus, GMM may be more useful than BSLMM if the true size of each **(*β***_***m***_)_***j***_ is related to the size of the corresponding **(*α***_***a***_)_*j*_, and vice-versa.

### Group 2 Methods

This group of methods directly estimate the global indirect effect without producing estimates of its mediator-specific contributions.

#### PCMA

Principal component mediation analysis (PCMA), proposed by Huan and Pan (2016), was an early method for multiple-mediator mediation using principal component analysis (PCA)^39^. The authors perform PCA on the residual matrix of the mediator models, then use the *p* by *r* loading matrix ***Q*** to transform the matrix ***M*** into a new set of mediators, ***M***\*, which are uncorrelated conditional on *A* and ***C***. The transformed mediators then replace the original mediators in the analysis, and because they are uncorrelated, the outcome and mediator models can be fit without issue. Although the mediators have been transformed, and the mediator-specific contributions **(*α***_***a***_)_*j*_**(*β***_***m***_)_*j*_ no longer correspond to the original *j*^th^ mediator, the global indirect effect ***α***_***a***_^*T*^***β***_***m***_ can still be estimated with its original interpretation. The authors set *r* to equal *p*, though this is only possible if *p* is less than *n*.

#### SPCMA

Zhao et al (2019) proposed sparse principal component analysis (SPCMA) to improve the interpretability of the results from PCMA^40^. In PCMA, the transformed mediators are difficult to interpret because they are sums of all *p* original mediators; whereas in SPCMA, the loading matrix ***Q*** is sparsified, meaning that each transformed mediator is only a sum of a few of the original mediators. The results are easier to interpret because, if a specific transformed mediator has a large effect, it can potentially be traced back to the original mediators which were used to construct it. SPCMA induces bias in its estimation compared to PCMA, but it can be helpful for identifying groups of mediators which may be active.

#### HILMA

High-dimensional linear mediation analysis (HILMA), proposed by Zhou (2020), estimates ***α***_***a***_^*T*^***β***_***m***_ with a complex, de-biased penalized regression approach^38^. The mathematics of the procedure are beyond the scope of this text, but the proposed estimator has asymptotic properties for testing whether ***α***_***a***_^*T*^***β***_***m***_ is zero, and can also be applied when there are multiple (but not high-dimensional) exposures.

### Group 3 Methods

The last group of methods is fundamentally distinct from the others: Instead of fitting the original mediation models (Group 1), or estimating the mediation effect without fitting the models (Group 2), they reconceptualize the causal structure of the problem to produce results with unique interpretations. Like any method, they should only be applied when their assumptions about the causal structure are reasonable.

#### HDMM

High-dimensional multivariate mediation (HDMM), proposed by Chén et al. (2018), is similar to PCMA in that it uses dimension reduction, but chooses the loading vectors with a likelihood-based approach instead of PCA^41^. The loading vectors are referred to as “directions of mediation,” each vector specifying a linear combination of mediators which contribute to the likelihood of the mediation models. Hence, HDMM implicitly assumes that there are latent, unmeasured mediating variables that can be represented as linear combinations of the observed mediators. The results of HDMM are difficult to interpret, but it can still be useful for identifying whether there is any mediation through ***M*** at all, and for identifying large subsets of mediators that contribute to that mediation.

#### LVMA

Latent variable mediation analysis (LVMA), proposed by Derkach et al. (2019), assumes that ***M*** itself is not involved in mediation, but rather, that there are a small number of unmeasured mediators, ***F***, which transmit the effect of *A* to *Y* and which also cause changes in ***M***^43^. In other words, LVMA assumes explicitly what HDMM assumes implicitly, and the results of the two methods have a similar structure. A key feature of LVMA is that the ***F*** → ***M*** associations are sparsified, meaning that the method can be used for detecting relevant mediators in ***M***. An observed mediator would be considered active if it is associated with a latent mediator that is itself associated with *A* and *Y*.

### Simulation study

#### Simulation settings

We evaluate the above methods with a simulation study. To contrast them under diverse conditions, we consider three different settings of mediation: (1) a baseline setting in which the mediation signals are sparse and the (potential) mediators are moderately correlated, (2) a high-correlation setting with sparse signals, and (3) a moderate correlation setting in which the signals are non-sparse. Within each of these settings, we also vary the degree of mediation by modifying three parameters: the proportion of variance in ***M*** that is explained by *A* among those associated with *A* (PVE_A_), the proportion of the variance of *Y* that is explained by the direct effect (PVE_DE_), and the proportion of the variance of *Y* that is explained by the global indirect effect (PVE_IE_). For a baseline case, we let PVE_A_ equal 0.20 and PVE_DE_ and PVE_IE_ both equal 0.10; then, in three additional cases, we sequentially decrease one of these parameters by half, weakening the signal, and set the other two parameters to their values from the baseline. Between Settings (1) to (3), this amounted to 12 unique data-generating mechanisms in total. Each of these was evaluated with a sample size of 1,000 and 2,500, with the number of potential mediators fixed at 2,000. All combinations of settings are listed below in Table 5.

**Table 5.** Complete list of settings in simulation study.

| Number of potential mediators ($p$) | Sample Size ($n$) | Sparsity of signals | Degree of correlation | PVE<sub>A</sub> | PVE<sub>IE</sub> | PVE<sub>DE</sub> |
| --- | --- | --- | --- | --- | --- | --- |
| 2000 | 2500 | Sparse | Baseline | 0.20 | 0.10 | 0.10 |
| 2000 | 2500 | Sparse | Baseline | 0.20 | 0.05 | 0.10 |
| 2000 | 2500 | Sparse | Baseline | 0.10 | 0.10 | 0.10 |
| 2000 | 2500 | Sparse | Baseline | 0.20 | 0.10 | 0.05 |
| 2000 | 2500 | Sparse | High | 0.20 | 0.10 | 0.10 |
| 2000 | 2500 | Sparse | High | 0.20 | 0.05 | 0.10 |
| 2000 | 2500 | Sparse | High | 0.10 | 0.10 | 0.10 |
| 2000 | 2500 | Sparse | High | 0.20 | 0.10 | 0.05 |
| 2000 | 2500 | Non-sparse | Baseline | 0.20 | 0.10 | 0.10 |
| 2000 | 2500 | Non-sparse | Baseline | 0.20 | 0.05 | 0.10 |
| 2000 | 2500 | Non-sparse | Baseline | 0.10 | 0.10 | 0.10 |
| 2000 | 2500 | Non-sparse | Baseline | 0.20 | 0.10 | 0.05 |
| 2000 | 1000 | Sparse | Baseline | 0.20 | 0.10 | 0.10 |
| 2000 | 1000 | Sparse | Baseline | 0.20 | 0.05 | 0.10 |
| 2000 | 1000 | Sparse | Baseline | 0.10 | 0.10 | 0.10 |
| 2000 | 1000 | Sparse | Baseline | 0.20 | 0.10 | 0.05 |
| 2000 | 1000 | Sparse | High | 0.20 | 0.10 | 0.10 |
| 2000 | 1000 | Sparse | High | 0.20 | 0.05 | 0.10 |
| 2000 | 1000 | Sparse | High | 0.10 | 0.10 | 0.10 |
| 2000 | 1000 | Sparse | High | 0.20 | 0.10 | 0.05 |
| 2000 | 1000 | Non-sparse | Baseline | 0.20 | 0.10 | 0.10 |
| 2000 | 1000 | Non-sparse | Baseline | 0.20 | 0.05 | 0.10 |
| 2000 | 1000 | Non-sparse | Baseline | 0.10 | 0.10 | 0.10 |
| 2000 | 1000 | Non-sparse | Baseline | 0.20 | 0.10 | 0.05 |

#### Simulated dataset creation

First, to obtain sparse mediation effects for Settings (1) and (2), we assume that 1,920 of the 2,000 coefficients **(*α***_***a***_)_*j*_ and **(*β***_***m***_)_*j*_ are zero and the remaining 80 are standard normal. Twenty of the nonzero **(*α***_***a***_)_*j*_ and **(*β***_***m***_)_*j*_ are chosen to overlap and have **(*α***_***a***_)_*j*_**(*β***_***m***_)_*j*_ not equal to zero. To obtain non-sparse signals for Setting (3), we sample the previously zero coefficients from a normal distribution with mean zero and standard deviation 0.2. (These parameter vectors are sampled only once, at the start of the simulations, so that the global mediation effect is held constant, but we shuffle the mediators in each dataset so that different mediators are assigned the effects each time.) Once we have these, we obtain a single simulated dataset by sampling *A*_*i*_ from a standard normal distribution, then produce ***M***_***i***_ from model (4) assuming there are no covariates. We add noise to ***M***_***i***_ by sampling residuals from a multivariate normal distribution with mean **0**_***p***_ and variance **Σ**, where **Σ** is derived by shuffling and then tuning the variance-covariance of the observed methylation data (see supplementary section 1). In Settings (1) and (3), we tune **Σ** so that the correlations between mediators range from -0.37 to 0.49, and in Setting (2), so that they range from -0.58 to 0.75. We fix PVE_A_ by scaling **Σ** appropriately based on ***α***_***a***_. Finally, we define *Y*_*i*_ based on model (3) assuming the residuals are Normal(0,σ^2^), choosing *β*_*a*_ and σ^2^ to yield the desired PVE_DE_ and PVE_IE_.

#### Evaluation

We evaluate the methods by applying them to 100 replicates of each setting in Table 5. We omit SPCMA, GMM, and LVMA for computational reasons, as they are too computationally costly to deploy on so many replicates, and omit HDMM because it does not have an estimand that is comparable to the others. We include a one-at-a-time approach—in which the mediator are assessed individually using traditional mediation analysis and the joint significance test^44^—as a baseline for comparison. When running HIMA, HDMA, MedFix, and pathway LASSO, we pre-screen the mediators to only include the *n*/log(*n*) mediators with the strongest associations with *Y* adjusting for *A*, which is the approach recommended by the HIMA and HDMA authors^34,35^ (see supplementary section 2 for more details). For comparison metrics, we use (1) the true positive rate for detecting active mediators,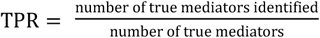; (2) the mean squared error in estimating the mediation contributions of inactive mediators, 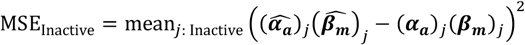; (3) the mean squared error in estimating the mediation contributions of active mediators,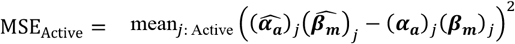 ; and (4) the percent relative bias in estimating the global indirect effect, 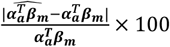. In the non-sparse setting, since all the mediators contribute to the indirect effect, we consider the “active” ones to be those whose mediator-outcome and exposure-mediator effects both come from the distribution with higher-variance, and the others inactive. Each metric is computed for each dataset to the applicable methods, and we report the average and a 95% empirical confidence interval over the 100 replicates.

#### Scalability comparison

We compare the scalability of the methods by assessing their processing time on simulated datasets of two sizes: one with 100 observations and 200 mediators and one with 1,000 observations and 2,000 mediators. For the larger dataset, we use one of the datasets created for the simulation study, and for the smaller dataset, we subset the rows and columns of ***M*** and the entries in ***A*** and ***Y***. Run times are assessed on a single core of an Intel(R) Xeon(R) Gold 6242R CPU @ 3.10GHz processor. We attempt each method 30 times and report the mean and interquartile range of the computation times. Since SCPMA and BSLMM tend to be time-consuming, we approximate their run times by downscaling the appropriate parameters: In particular, since the desired number of principal components in SPCMA is 100, we use only 2 principal components and scale the computing time by 50; and since the desired number of posterior samples in BSLMM is 30,000, we draw only 750 samples and scale the result by 40. Ad hoc experimentation confirmed that the methods were approximately linear with respect to these inputs.

### Data application with MESA

To demonstrate how these methods can be applied to observed DNAm data, we evaluate the association between SES and HbA1c and its potential mediation through DNAm. For the exposure, we use a binary variable that indicates low educational attainment (less than a 4-year college degree); for the outcome, we use HbA1c, a continuous variable that reflects average three-month blood glucose level. Our data for this portion come from the Multi-Ethnic Study of Atherosclerosis (MESA), a US population-based longitudinal study^17^. Out of 6,814 total participants, a random subsample of 1,264 had their DNAm measured at 484,882 CpG sites. We limit our analysis to the 963 participants who (1) had methylation data, (2) had no missing data for the required variables, (3) consented to genetic and phenotypic use through the database of Genotypes and Phenotypes (dbGaP) (phs000209.v13.p3), and (4) were not on diabetes medication, which can cause changes in HbA1c (Fig. 6). Standard quality control filters reduced the number of CpG sites to 402,339. Since it is not statistically or computationally feasible to include so many mediators at once, we used a screening procedure to reduce that number further, fitting model (6) below for each mediator separately to choose the 2,000 CpG sites at which DNAm was most strongly associated with education based on the **(*α***_***a***_)_*j*_ p-value. These 2,000 formed the baseline set of CpGs for our analysis. DNAm was measured using M-values, defined as the log-2 ratio of the methylated to unmethylated probe intensities, which has the advantage of occurring on a continuous and unbounded scale^56^. For more details see supplementary section 3. A model for the proposed mechanism is given by

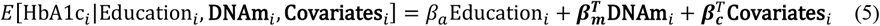

and

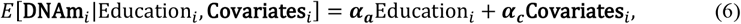

where the covariates include age, sex, race, and the estimated proportions of residual non-monocytes (i.e., neutrophils, B cells, T cells, and natural killer cells) as fixed effects and methylation chip and position as random effects.

**Fig. 6.**
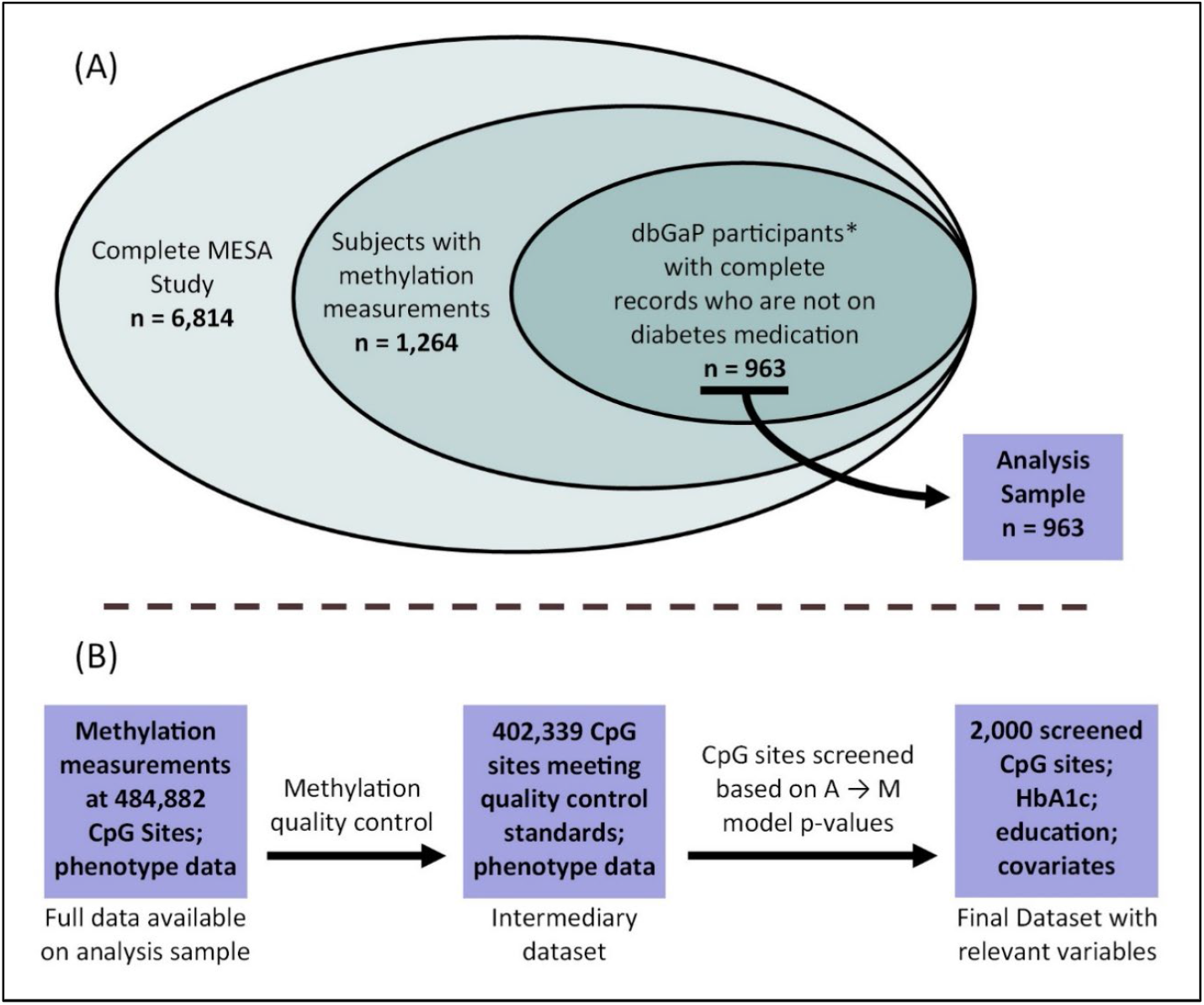
Pre-processing of MESA methylation data. *Participants who consent to genetic and phenotypic data use, and whose data is available on dbGaP.

We performed mediation analysis on the final dataset of 963 individuals and 2,000 CpG sites. All of the mediation methods described above were included except for GMM and LVMA, which again were too costly computationally. Although it is reasonable for some of the methods to include all 2,000 CpG sites directly in the multivariable model, HIMA and HDMA involve sure independence screening^57^ to reduce the number of mediators in advance to *n*/log(*n*), where *n* is the sample size. For the sake of consistency across the penalized regression methods, we do so with not only HIMA and HDMA, but also MedFix and pathway LASSO, including only the 141 (963/log(963)) CpG sites most associated with low education (a direct extension of the initial screening). (Note that, for HIMA and HDMA, this screening is *part* of the proposed method, not separate from it, but for MedFix and pathway LASSO the additional screening is still beneficial for the sake of comparing methods and for statistical and computational efficiency). Additional pre-screening is not necessary for PCMA, SPCMA, BSLMM, and HILMA, and we include all 2,000 CpG sites directly; however, in HDMM, which cannot accommodate *p* > *n* simplistically, we again use only twice-screened subset of 141 sites. For the sake of comparison with multivariate methods, we also include a one-at-a-time mediation method based on linear regression and the joint significance test. For estimating the total effect, the methods PCMA, SPCMA, BSLMM, and Pathway LASSO all produce estimates of the direct effect, so we can estimate the total effect by summing the estimated direct and global indirect effects. Since the methods HIMA, HDMA, and MedFix do not produce estimates of the direct effect, we first estimate the total effect on its own by fitting model (5) with the mediators excluded, then subtract the estimated global indirect effect from this value to obtain an estimate of the direct effect. As none of the high-dimensional methods are built to directly handle random effects as covariates, we regress these out of the outcome variable and potential mediators in advance. For the fixed effect covariates, HIMA, HDMA, MedFix, and BSLMM allow one to include them directly; whereas in PCMA, SPCMA, HILMA, HDMM, and pathway LASSO, we regressed them out in advance from the outcome and mediators. Continuous variables (including HbA1c and the mediators) were standardized for all methods. All analysis was conducted using R version 4.2.1.

## Data Availability

Code for generating the simulation data can be found at https://github.com/dclarkboucher/mediation_DNAm. Data used in the DNAm analysis can be obtained through the MESA Data Coordinating Center (https://www.mesanhlbi.org/).

https://www.mesanhlbi.org/

## Data Availability

Data used for the simulation study are available from the authors upon request. Data used in the DNAm analysis can be obtained through the MESA Data Coordinating Center (https://www.mesanhlbi.org/).

## Code Availability

R scripts for the analysis are available at https://github.com/dclarkboucher/mediation_DNAm. Our R package “hdmed” can be found at https://github.com/dclarkboucher/hdmed.

## Acknowledgements

MESA and the MESA SHARe project are conducted and supported by the National Heart, Lung, and Blood Institute (NHLBI) in collaboration with MESA investigators. Support for MESA is provided by contracts 75N92020D00001, HHSN268201500003I, N01-HC-95159, 75N92020D00005, N01-HC-95160, 75N92020D00002, N01-HC-95161, 75N92020D00003, N01-HC-95162, 75N92020D00006, N01-HC 95163, 75N92020D00004, N01-HC-95164, 75N92020D00007, N01-HC-95165, N01-HC-95166, N01-HC-95167, N01-HC-95168, N01-HC-95169, UL1-TR-000040, UL1-TR-001079, UL1-TR-001420, UL1-TR-001881, and DK063491. The MESA Epigenomics & Transcriptomics Studies were funded by NIH grants 1R01HL101250, 1RF1AG054474, R01HL126477, R01DK101921, and R01HL135009. Co-authors of this manuscripts were partially supported by NHLBI grant R01HL141292, NSF grant DMS1712933, and NIH grants R01HG008773 and 1UG3CA267907.

