## Supplementary Information for "Methods for Mediation Analysis with High-Dimensional DNA Methylation Data: Possible Choices and Comparison"

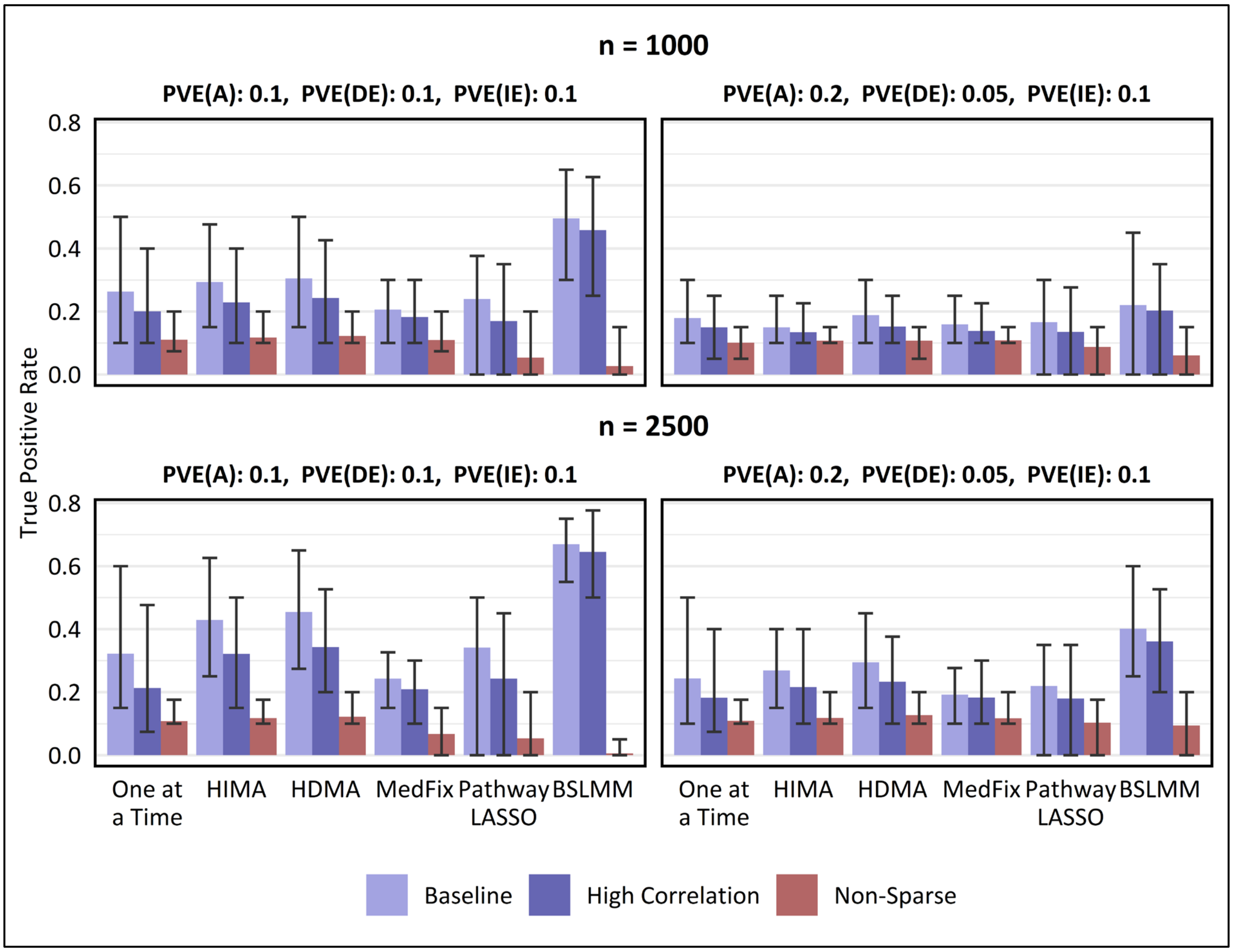


**Supplementary Fig. 1.** Mean true positive (TPR) rate and 95% empirical confidence interval for detecting active mediators in 100 simulated datasets. In the baseline and high-correlation-among-mediators settings, TPR is for distinguishing mediators which contribute to the global mediation effect from those which do not, whereas in the non-sparse setting, where all mediators contribute, TPR is for distinguishing mediators whose contributions were sampled from a high-variance distribution from those whose contributions were sampled from a low-variance distribution. False discovery rate was capped below 10% by a proper choice of the p-value threshold (one-at-a-time, HIMA, HDMA, MedFix), posterior inclusion probability threshold (BSLMM), or method tuning parameter (P-LASSO).

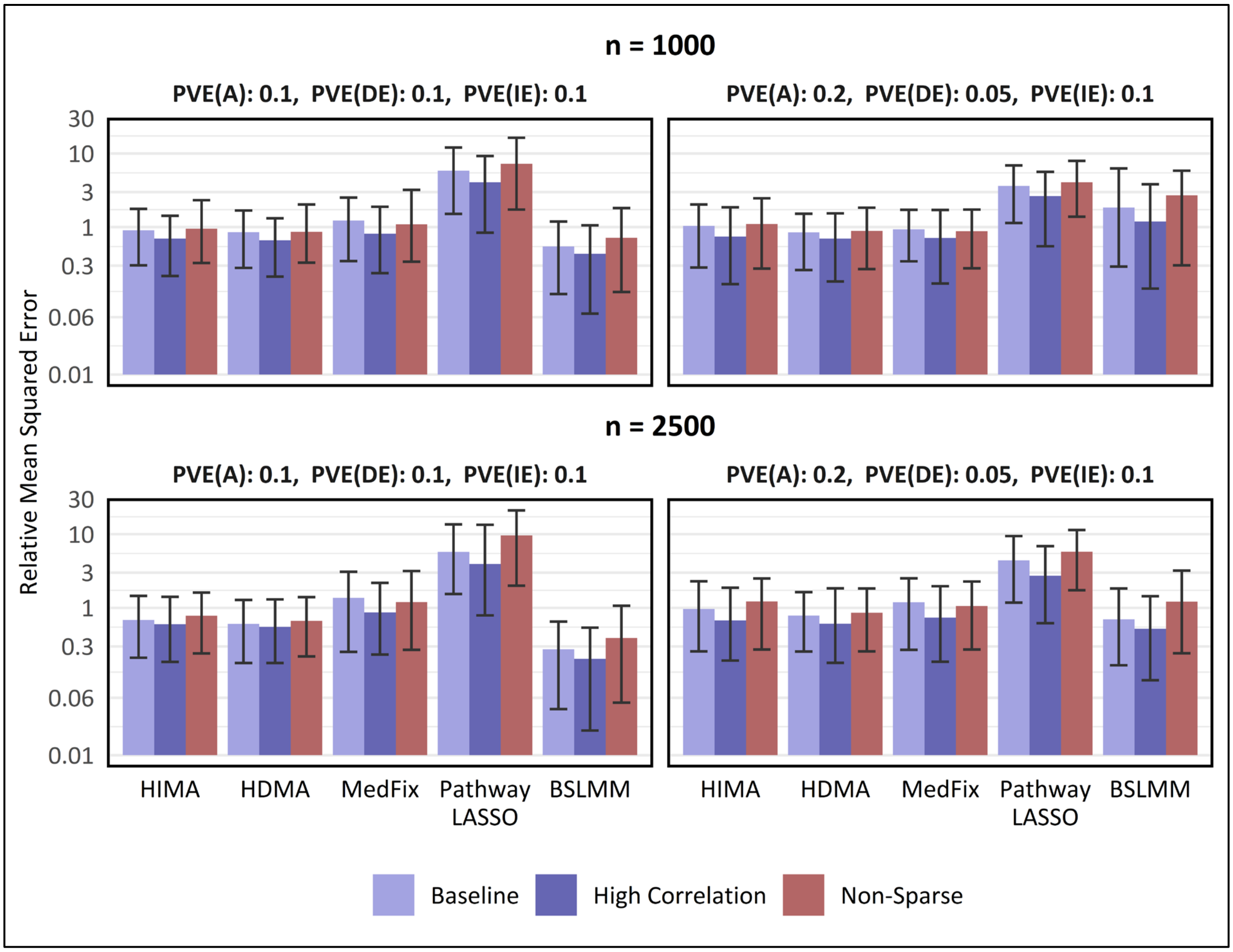


**Supplementary Fig. 2.** Mean relative mean squared error (rMSE) and 95% empirical confidence interval for estimating mediation contributions among active mediators in 100 simulated datasets, relative to the one mediator at a time method. Y-axis is on a $\log_{10}$ scale. For baseline and high-correlation-between-mediators settings, active mediators which contribute to the global mediation effect, whereas in the non-sparse setting, where all mediators have some contribution, active mediators are those whose contributions were sampled from a distribution with large variance instead of small.

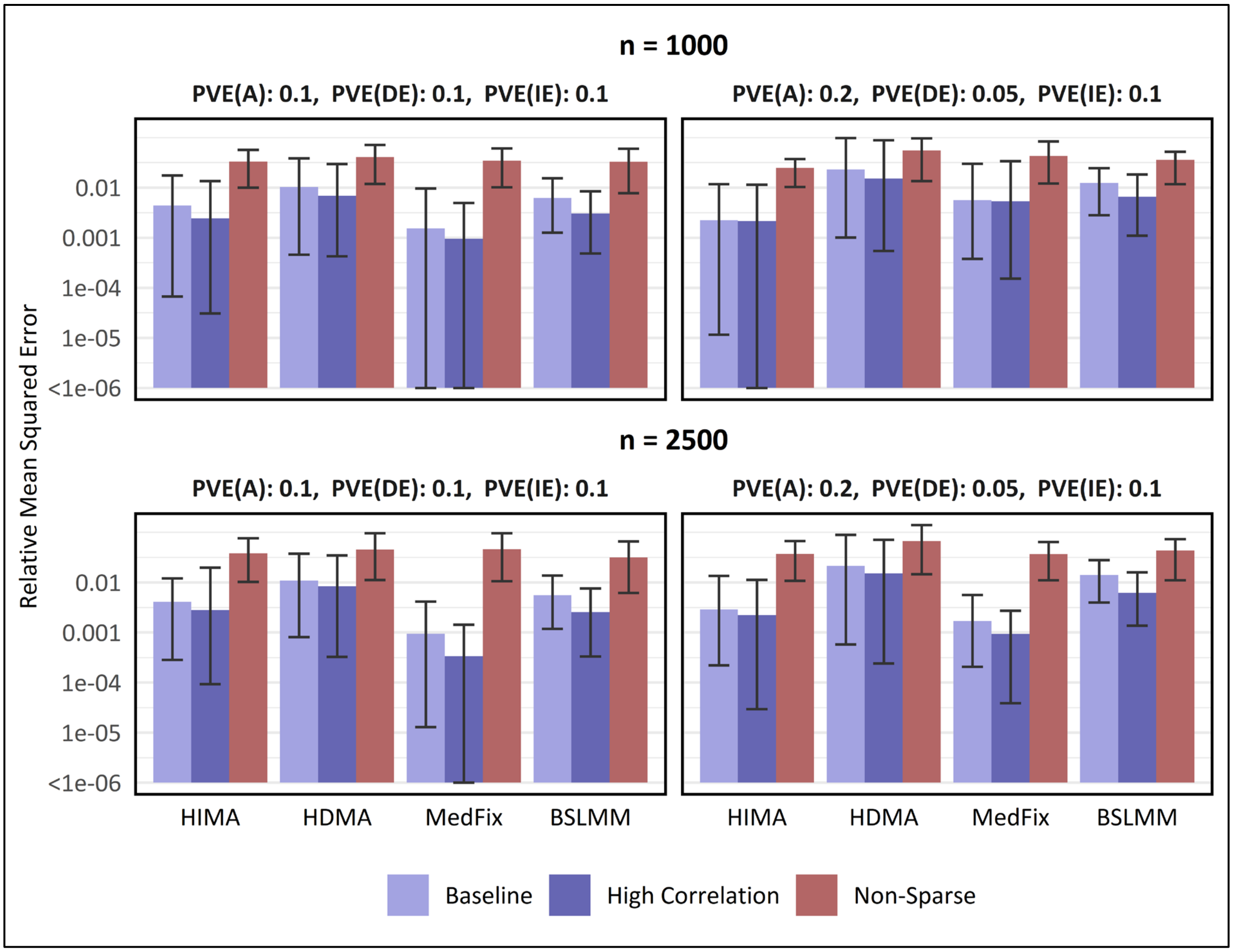


**Supplementary Fig. 3.** Mean relative mean squared error (rMSE) and 95% empirical confidence interval for estimating mediation contributions among inactive mediators in 100 simulated datasets, relative to the one mediator at a time method. Y-axis is on a $\log_{10}$ scale. For baseline and high-correlation-between-mediators settings, inactive mediators are those which have no mediation contribution, whereas in the non-sparse setting, where all mediators have some contribution, inactive mediators are those whose contributions were sampled from a distribution with small variance instead of large.

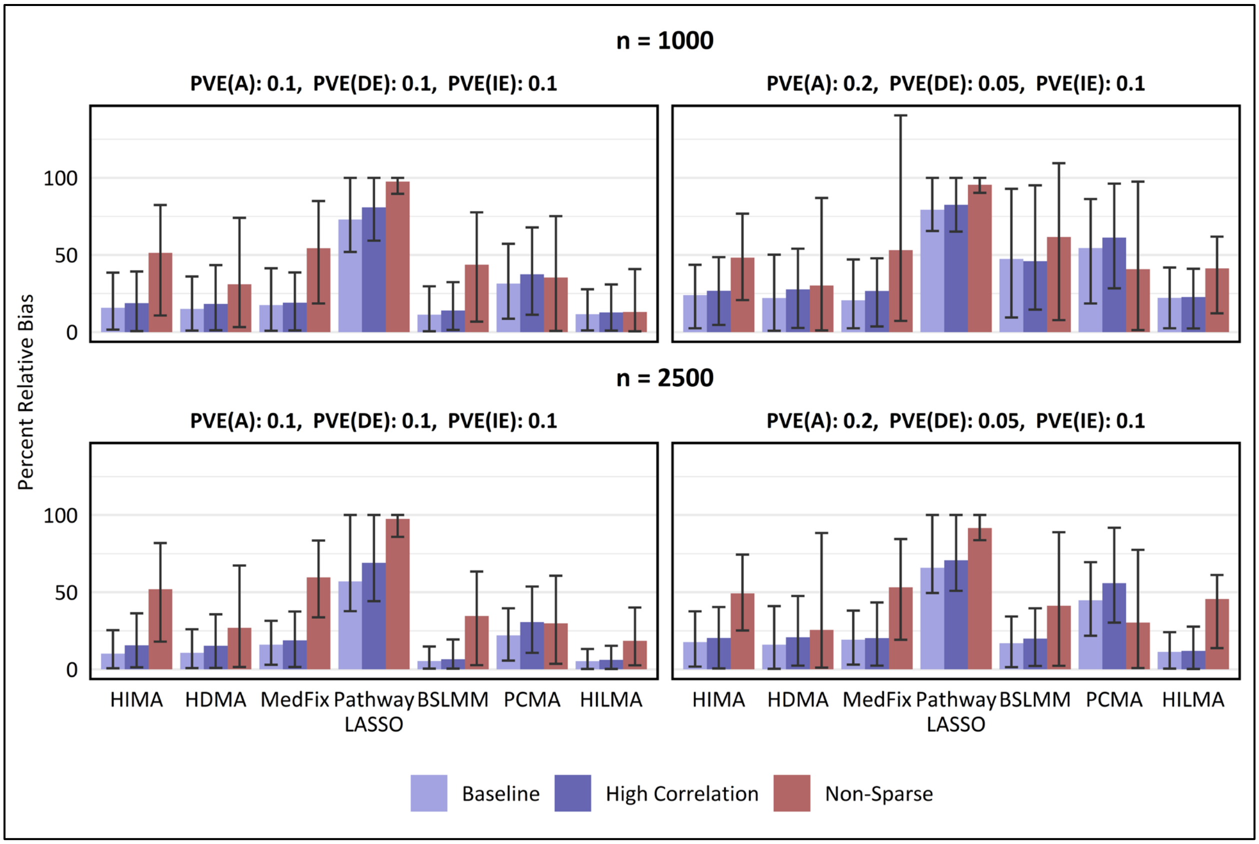


**Supplementary Fig. 4.** Mean percentage relative bias in estimating the global mediation effect across 100 simulated data replicates, with intervals representing the inner 95% range.

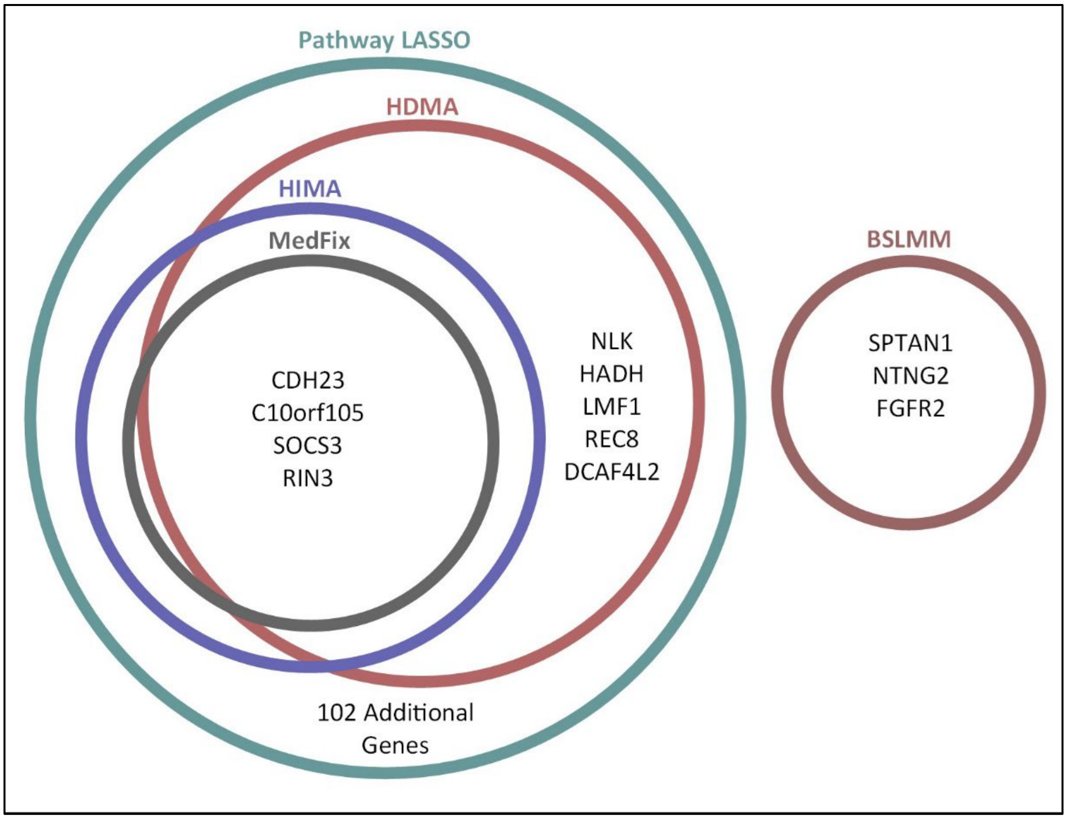


**Supplementary Fig. 5.** Genes containing or near CpG sites selected as active mediators between low socioeconomic status and HbA1c by methods for high-dimensional mediation analysis. CpG sites were linked to genes using R Bioconductor package "IlluminaHumanMethylation450kanno.ilmn12.hg19". Additional genes detected by Pathway LASSO listed in Supplementary File 1.

**Supplementary Table 1. Memory usage of mediation methods by dataset dimensions**

| Method | RAM Usage (Megabytes) | |
|---|---|---|
| | $n$ = 100, $p$ = 200 | $n$ = 1,000, $p$ = 2,000 |
| BSLMM | 114 | 484 |
| HDMA | 238 | 342 |
| HIMA | 233 | 306 |
| MEDFIX | 192 | 286 |
| PCMA | 315 | 638 |
| PLASSO | 72 | 115 |
| SPCMA | 378 | 1,222 |
| HDMM | 359 | 687 |
| HILMA | 106 | 603 |

### Section 1. Tuning of variance covariance matrix in simulations

As described in the main text, the residuals of the mediator models are sampled from a multivariate normal distribution with mean $\mathbf{0}_p$ and variance $\boldsymbol{\Sigma}$. To obtain $\boldsymbol{\Sigma}$, we begin by computing the true residual variance-covariance matrix from the real data analysis with DNAm. This matrix is singular due to the fact that there are more CpG sites (2,000) being analyzed than observations (963). To make the matrix non-singular, we first standardize it so that each mediator has residual variance 1, then add a small penalty, $r$, to every element of the diagonal. Then we standardize a second time. This procedure is necessary because if we do not add such a penalty, the variance-covariance matrix remains non-invertible, so the residuals once sampled will be linearly dependent regardless of the chosen sample size. Moreover, since the size of $r$ determines the degree of correlations between mediators—larger values resulting in weaker correlations—we can change the correlations between simulation settings by tweaking $r$. For the baseline correlation level, we set $r$ to be 1, forcing the correlations to range from -0.37 to 0.49, and for the high-correlation setting, we set $r$ to be 0.5, letting the correlations range from -0.58 to 0.75.

### Section 2. Method application details

One-at-a-time

We assess the mediators "one at a time" using the standard mediation models proposed by Baron and Kenny (1986)[1]. These are analogous to the models established in the main text (i.e., models 1 and 2) except that they treat each mediator separately. We assess the significance of each mediator using the max-P test (or joint significance test), in which the maximum of the exposure-mediator and mediator-outcome association p-values is tested against the significance level[2]. In the simulated data analysis, we identify active mediators by thresholding these max p-values so that at most 10% of the "significant" mediators are not true mediators, ensuring that the false discovery proportion (FDP) on that dataset is below 0.10 and the false discovery *rate* (FDR) across all datasets is also below 0.10. In the real data analysis, we use a linear mixed model instead of linear regression so that we can include the appropriate confounding variables as random effects; in particular, age, sex, race, and the estimated proportions of residual non-monocytes are adjusted for as fixed effects and methylation chip and position as random effects (to address potential batch effects). We fit this model with the "lmerTest" package in R[3]. (The mediation model from this set up is the same model used to screen the CpG sites from 402,339 down to 2,000.) To identify CpG sites involved in mediation, we take the max-P values from these models and adjust them using the "qvalue" function from the "qvalue" package[4], then compare them to a signficance level of 10% to control the FDR.

HIMA

We apply HIMA directly using the "hima" function from the "HIMA" package in R. On the simulated datasets, we apply sure independence screening[5] as recommended by the HIMA authors[6] to reduce the set of mediators to the $n/\log(n)$ mediators mostly strong associated with $Y$, adjusting for $A$ (screening based on the outcome model). The number $n/\log(n)$ is chosen to encourage dimension reduction while maintaining the accuracy of the screening procedure[5]. For FDR correction, we choose a p-value threshold for each dataset to ensure that the false discovery proportion for that dataset is below 0.10.

On the DNAm data from MESA, so that the sure indepdent screening matches the original screening used to arrive at 2,000 CpG sites, we screen on the association of each CpG site with the exposure, low education, using the same linear mixed model used in the mediator model of the one-at-a-time approach. (The HIMA and HDMA authors suggest that screening based on either the outcome model or the mediator model is acceptable[6,7]). We identify CpGs as noteworthy based on whether their estimated mediation contribution is not zero. Although HIMA does produce p-values (which we use for thresholding in the simulated data analysis), the p-values are based on the subsequent fitting of an ordinary linear regression after the penalized model has performed feature selection; hence, they can be expected to be overconfident. It is for this reason that throughout our DNAm analysis, we avoid commenting on the "statistical significance" of the mediation contributions and focus only on identifying which ones are "noteworthy".

HDMA

We apply HDMA using the "hdma" function provided by Gao et al. at their repository https://github.com/YuzhaoGao/High-dimensional-mediation-analysis-R/blob/master/HDMA.R. Application of the "hdma" function is similar to application of the "hima" function from the "HIMA" package, and we apply HDMA identically to how we apply HIMA as described above.

MedFix

Although code for MedFix is provided in the GitHub repository located at https://github.com/QiZhangStat/highMed, it is designed for the setting where both $A$ and $M$ are high-dimensional (the primary setting for which MedFix is intended). We provide code for implementing MedFix for a single exposure in our package "MultiMed." Although MedFix does not explicitly involve sure independence screening like HIMA or HDMA, we elect to use screening for MedFix as well so that the penalized regression methods (which are very similar) are all applied in the same systematic fashion. Other details on how MedFix was implemented are comparable to HIMA as described above.

PCMA

We apply PCMA using the "mcma_PCA" function in the "SPCMA" R package. For both the simulated data analysis and DNAm data analysis, we set the number of principal components to be 100. Although it may be preferable in some cases to choose this number to be larger, capturing more of the variance of $M$, it is better for the sake of our comparison to use the same number of principal components in both PCMA and SPCMA, and applying SPCMA with more than 100 principal components would be extremely computationally costly. Further, in the simulated data analysis where SPCMA was not used at all, choosing the maximal number of principal components (the minimum of $n$ and $p$) resulted in extreme variability in the estimated total indirect effect and did not cause the method to perform better than it did with 100.

SPCMA

We apply SPCMA using the "spcma" function from the "spcma" package. Analogously to PCMA, we set the number of sparse principal components to be 100. To create sparsity in the principal component loading vectors, SPCMA uses the fused LASSO penalty[8], which in addition to the L1 penalty used by the regular LASSO, penalizes the difference in coefficient effects between adjacent variables, inducing smoothness. The parameter $\gamma$ represents the ratio of the L1 penalty to the fusion penalty, and can be chosen in advance. We set $\gamma$ to be 2 so that the L1 penalty is emphasized, as "adjacent" CpG sites could still be far apart in the genome and should not be expected to have similar effects, making the fusion penalty unimportant. However, different choices of $\gamma$ did not appear to dramatically change our results. We test for the significance of the 100 transformed mediators using the bootstrapping method proposed by the SPCMA authors, with 100 bootstrap samples and bias-corrected confidence intervals[9].

Pathway LASSO

Code for implementing pathway LASSO is provided by the authors in the GitHub repository located at https://github.com/zhaoyi1026/PathwayLasso. Like MedFix, pathway LASSO does not explicitly involve pre-screening the mediators, but we still conduct the pre-screening procedure from HIMA and HDMA so that the penalized regression methods HIMA, HDMA, MedFix, and pathway LASSO are more comparable. This is also beneficial computationally, as it is slow to apply pathway LASSO directly to data with 2,000 mediators and 2,500 or 1,000 observations. Another aspect of pathway LASSO implementation is selecting the tuning parameters. These include $\omega$, which controls a LASSO-like penalty on each $(\boldsymbol{\alpha_a})_j$ and $(\boldsymbol{\beta_m})_j$, $\phi$, a convexity parameter, and $\lambda$, which controls a complex penalty function including the product terms $(\boldsymbol{\alpha_a})_j(\boldsymbol{\beta_m})_j$. For our analysis, $\phi$ is fixed at 2, as it is in fMRI study presented by

Zhao and Luo (2022), who show that pathway LASSO is not sensitive to the choice of this parameter[10]. For the other parameters, we fix the ratio of $\omega$ to $\lambda$ to be 1 (i.e., forcing the parameters to be equal), as this ratio performed best in the Zhao and Luo's simulation study. We then attempt pathway LASSO using 45 different values of $\lambda$. In the simulated data analysis, we choose the optimal value of $\lambda$ based on the observed false discovery proportion, selecting the smallest $\lambda$ for which fewer than 10% of the selected mediators are true mediators in that simulated dataset. This is necessary because pathway LASSO does not provide a method to test the statistical significance of the effects using p-values, like in HDMA, HIMA, and MedFix, preventing us from using p-values for thresholding. In the DNAm analysis, we use the variable selection stability criterion suggested by the authors[11] with the code provided in the GitHub package. Like in the other penalized regression methods, CpG sites are considered noteworthy if their estimated mediation contribution is not zero.

BSLMM

We apply BSLMM using the "bama" function from the "bama" package in R[12]. Application of BSLMM depends on several parameters: *lm0*, *lm1*, *lma1*, *l*, and *k*. These are, respectively, the scale parameter for the inverse-gamma prior for the small-variance $(\boldsymbol{\alpha_a})_j$ and $(\boldsymbol{\beta_m})_j$, the scale parameter for the inverse-gamma prior for the large-variance $(\boldsymbol{\beta_m})_j$, the scale parameter for the inverse-gamma prior for the large-variance $(\boldsymbol{\alpha_a})_j$, the scale parameter for the inverse-gamma prior on the other coefficient variances, and the shape parameter for the inverse-gamma priors. In applying BSLMM, we fix *k* at 1 (the default setting) so that the prior mean of each variance is equal to the scale parameter. We also fix *l* at 1 (the default setting). In both the DNAm data analysis and simulated data analysis, we choose *lm1* by taking the variance of the absolute largest 10% of the marginal $(\boldsymbol{\beta_m})_j$ coefficients (i.e., the coefficient from the one-at-a-time method), and choose *lma1* similarly except with the marginal $(\boldsymbol{\alpha_a})_j$ coefficients. (When working on simulated data, we know the true variance of the coefficients exactly, but choosing these parameters based on the known truth is not possible in practice and therefore should be avoided in simulations to keep them fair. Part of the difficulty in applying BSLMM on real data is choosing these parameters appropriately, and the results of BSLMM tend to be sensitive to this choice).

HDMM

We apply HDMM using the "PDM_1" function from the "PDM" R package[13]. The "PDM_1" function computes weights for the first principal direction of mediation (PDM), which are then used to linearly combine the set of mediators into a single, transformed latent mediator. We analyze the transformed mediator using the "mediation" R package[14], with 2,000 Monte Carlo draws used for the quasi-Bayesian confidence intervals. HDMM cannot directly incorporate settings where $p$ is greater than $n$ when computing its PDMs, so we repeat our sure independence screening procedure from HIMA, HDMA, MedFix, and pathway LASSO prior to the analysis. (Note that HDMM *can* be applied when $p$ is greater than $n$, but it requires using population value decomposition[15]. However, population value decomposition is designed for longitudinal settings where each observation contains multiple measurements representing different time points. Our attempts to apply population value decomposition to our data were unsuccessful for this reason.)

HILMA

We apply HILMA using the “hilma” function from the “freebird” R package[16]. We set the tuning method to “uniform” rather than “AIC” as recommended by the HILMA authors, and we standardize the data prior to analysis. (For the simulated data analysis, we multiply the resulting total indirect effect by the standard error of Y to project the estimate back to the original scale).

### Section 3. MESA Study and Data Processing

Our data come from the Multi-Ethnic Study of Atherosclerosis (MESA), a United States population-based longitudinal study on the risk factors and progression of subclinical cardiovascular disease[17]. MESA recruitment began in July 2000 and lasted until August 2002, during which period 6,814 participants were recruited from study sites in Forsyth County, North Carolina; New York, New York; Baltimore County, Maryland; St. Paul, Minnesota; and Chicago, Illinois. Ages at recruitment ranged from 45 to 84 years. Multiple examinations since the beginning of the study captured data on clinical information, socio-demographic traits, lifestyle and behavioral characteristics, and other factors. A random subsample of 1,264 MESA participants were selected between April 2010 and February 2012 to have their DNAm measured using the Illumina Infinium HumanMethylation450 Beadchip on purified monocytes. Quality control filters reduced the number of CpG sites from 484,882 to 402,339; in particular, sites were removed if they had "detected" methylation levels in less than 90% of MESA samples at a p-value cutoff of 0.05, were within 10 base pairs of a single nucleotide polymorphism (SNP) based on Illumina annotation, had unreliable probes (i.e., had SNPs with minor allele frequency greater than 0.05 within 2 base pairs or cross reactive probes, recommended by DMRcate[18]), or overlapped with a repetitive element or region; while probes on sex chromosomes, SNPs, and other non-CpG targeting probes were not considered. The raw methylation measurements were transformed into M-values by taking the log-2 ratio of the methylated to unmethylated probe intensities. Further details are provided by Liu et al. (2013)[19].

Our outcome variable was HbA1c measured at Exam 5, which was standardized prior to the analysis. This provides a measure of the average three-month blood sugar level. For our exposure variable, low adult socioeconomic status, we use a binary indicator variable representing the lack of a 4-year college degree (1: less than a 4-year college degree, 0: otherwise). Since 402,339 CpG sites is too many to include in an analysis with only 963 samples, we screened CpG sites in advance to reduce that number further, including only, at most, the 2,000 CpG sites most strongly associated with the exposure. This association was measured for each site by the p-value of the education coefficient from a linear mixed model in which methylation was regressed onto education level (the binary indicator), age, sex, race, and the estimated proportions of residual non-monocytes as fixed effects, and methylation chip and position as random effects. This model is equivalent to the one-at-a-time mediation mediator model described above. For the methods which require or recommend additional screening, we do so with the same model, described in the main text and in supplementary section 2 above.
